# Mechanisms of Supportive Supervision and their effectiveness in Primary Health care of low and - and Middle-Income Countries: A systematic review

**DOI:** 10.1101/2025.10.09.25337702

**Authors:** Anda Nindi-Nyondo, Felix Chisoni, Thomas Mildestvest, Eivind Meland, Luckson Dullie, Eric Umar

## Abstract

**Background:** Supportive supervision (SS) is a systematic approach used to assess health facilities’ preparedness to provide quality healthcare service and support healthcare providers to identify and address barriers to the provision of quality health services. There is, however, inconclusive evidence of the full benefit and influence of SS on performance in different settings of primary health care (PHC). Additionally, not much is known about what constitutes an effective SS approach that can successfully improve service provision by influencing better problem solving. This review synthesised evidence on the effectiveness of mechanisms of SS in PHC in low- and middle-income countries (LMIC).

**Methods:** *Search strategy:* The first search was done in five relevant databases, then in the remaining database and finalised with a search in the reference list.

*Selection criteria:* Selection criteria were guided by population (primary health care providers), intervention (mechanism of supportive supervision) and outcomes (positive or negative) in the context of low- and middle-income countries. Selection was based initially on title and abstract, then full-text articles.

*Data extraction and analysis:* A data extraction tool was used to extract the relevant information to answer the review question. Studies were analysed through frequency counting of articles. And qualitative content analysis was used, and synthesised results were reported narratively.

**Results:** A total of 29 studies were identified, ranging from 2013 to 2023. Methodologies identified included 6 cluster randomised control trials, 15 quasi-experimental studies, 5 qualitative studies, 5 analytical cross-sectional studies and 3 cohort studies. The studies were from the following: Egypt, Ethiopia, Pakistan, India, Kenya, Zambia, Nigeria, Mozambique, Tanzania and Malawi.

Study results found multiple mechanisms used when conducting SS in PHC in LMIC. SS was either effective 69% (n=20), ineffective 3.5% (n=1) or had inconclusive 28% (n=8) results. The role of SS in primary care performance included improved attitudinal behaviour, knowledge, skills and practice, and health and patient outcomes.

**Conclusion:** There are multiple approaches to conducting SS, and the mechanisms yielded both positive, negative and inconclusive results, supporting evidence of inconsistencies in what constitutes an effective mechanism of SS. Multiple external factors such as incentive, training and transport logistics were seen to influence the role of SS in PHC.

## INTRODUCTION

Provision of quality health services in low- and middle-income countries is faced with numerous challenges, with the most common ones being shortage of staff compared to the high volume of patients most facilities receive; shortage of equipment and supplies due to misallocations and/or competing needs; lack of motivation among staff due to poor working conditions, especially for those in hard-to-reach rural areas; and lack of effective performance management systems that ensure good performance among health care providers (6). There are many interventions that are put in place to work on these health system challenges, with supportive supervision being one of the most used tools in multiple low- and middle-income countries (7).

Supportive supervision (SS) is a systematic approach used to assess health facilities’ preparedness to provide quality healthcare service, support healthcare providers to identify and address barriers to the provision of quality health services and ensure that the allocation of resources and services is done according to need (1). It is a process of helping staff to improve their knowledge, skills and work performance in a respectful and non-threatening manner (8). It focuses on monitoring performance towards set goals using data for decision-making and ensuring that services are provided according to the set standards.

There are different methods of providing supportive supervision that are used in primary health care facilities. The pattern tends to follow that a more experienced or higher-ranking or higher-qualified provider takes on the responsibility of providing oversight to someone in a lower-level facility. Since primary health care health professionals at times carry responsibilities that are above their level of training and/or experience (9) and are mostly manned by lower-level cadres in the health systems, supportive supervision is seen as a tool for ensuring performance according to the stipulated standard of practice.

This systematic review references the WHO 2008 module of supportive supervision as a gold standard on how supportive supervision is conducted. The WHO 2008 module looks at the following steps to approaching supportive supervision: 1) Setting up a supportive supervision system 2) planning regular supportive supervision visits 3) conducting a supervisory visit and 4) following up on activities. Setting up a supportive supervision system includes training a core set of supervisors, creating checklists and recording forms and ensuring appropriate resources are available. Planning regular supportive supervision visits includes scheduling the supervision mostly once a quarter. Conducting a supervisory visit which includes observation, use of data, problem-solving, on-the-job training and recording observation and feedback. And finally, follow up on agreed action items by the supervisor and supervisee, regular data analysis and feedback to all stakeholders.

The third and fourth steps are what this study considers as the approach to conducting SS. And from these 2 steps. The 6 components are considered as the components of supportive supervision. This includes: 1) Training the supervisor on how to conduct a supervision 2) Supervisors observing/monitoring supervisees using a checklist 3) The supervisor provides feedback to the supervisees. 4) The supervisor provides on-the-job training, mentoring and/or coaching in the areas that the supervisee falls short in. 5) Supervisor and supervisee developing an action plan on how to resolve the action items And 6) Reviewing of action items.

There is, however, not much known about what constitutes an effective SS approach that can successfully improve service provision by influencing better problem solving and better communication between the supervisor and supervisee (3–5). Additionally, there is inconclusive evidence of the full benefit and influence of supportive supervision on performance in different settings of primary health care (1,3,5). This systematic review synthesised evidence on the effectiveness of mechanisms of supportive supervision in primary health care in low- and middle-income countries. The following review questions were answered: 1) What are the publication characteristics of identified evidence? 2) Which supportive supervision mechanisms are used by primary health providers? 3) What are the outcomes of supportive supervision mechanisms in terms of effectiveness? What role is played by supportive supervision in primary care performance?

## 2. METHODS

### Design and Protocol registration

A systematic review design was followed according to JBI. The review protocol was developed and registered in PROSPERO in February 2024, registration number CRD42024498553. The reporting of this systematic review followed the guidelines stated by Preferred Reporting Items for Systematic Reviews and Meta-Analyses (PRISMA).

### Eligibility criteria

The inclusion and exclusion criteria were guided by population, intervention and outcomes in the context of primary healthcare and low- and middle-income countries (Table 1).

| Guiding characteristics | Inclusion criteria | Exclusion criteria |
| --- | --- | --- |
| Population | Health care workers involved in supportive supervision (either as supervisors or supervisees) in low- and middle-income facilities at primary health care levels were included in the | Health workers in the ward |
|  | study. This also included healthcare workers working in the community. |  |
| Intervention | Mechanisms of Supportive Supervision |  |
| Outcomes | Effective; ineffective; inconclusive |  |
| Context | Studies published in English describing the mechanism of supportive supervision in the primary health care level, both at the facility and community levels, were included in the review. | All studies looking at supportive supervision conducted in a ward environment were excluded in the study. |
|  | Low- and middle-income country | Outside LMIC |
| Type of studies | All qualitative, quantitative or mixed-method studies describing mechanisms of supportive supervision in primary health care in low- and middle-income countries were included in the studies. | Letters, correspondence, position papers, conference papers and unpublished data were excluded in the review. Publications unavailable in English were excluded. A study was also excluded if the full text was unavailable from online databases. |
| Timeline | The studies between 2013 to 2023 were included in the study for review. | Articles published before 2013 |

### Information sources and search

A literature search was conducted from 12th to 16th February 2024 across the following databases: PubMed, Scopus, Embase, and Web of Science by the librarian (FC). The following search terms were used: “Supportive supervision”, “supervision mechanisms”, “supervision models”, “mentorship”, “supervision models”, “primary healthcare”, “primary level health facility”, “health care providers”. Search was restricted to low- and middle-income countries. Initially, articles were searched in two relevant databases, namely PubMed and Scopus, followed by the remaining ones (Embase and Web of Science) and finally the reference lists. The exact search string used for each database is available in Appendix 2.

### Study selection

Prior to the selection process, duplicates were removed. Two reviewers (ANN and FC) independently screened the retrieved articles based on their titles and abstracts and full texts using a Google Spreadsheet form. At each stage, any disagreements were resolved through discussion and consensus between AN and FC, with input from a third reviewer (LWD) as needed.

### Data extraction

Data extraction was conducted by ANN and reviewed by FC. As informed by the specific review questions, ANN extracted data under the following subheadings: author and year of publication, study design, health domain, study setting, mechanism of supportive supervision, the effectiveness and the role of supportive supervision. FC verified the extracted data based on the subheading categories.

### Risk of bias assessment

To attain data that is accurate and of good quality, the two independent reviewers (ANN and FC) went through the steps of the review independently (Bailey et al., 2015). However, due to limited resources for the study, the same reviewers who extracted the data cross-checked the data extracted to ensure that there were no errors. And the Joanna Briggs Institute (JBI), 2020 critical appraisal checklist was used for quality assessment.

### Data analysis and synthesis

Evidence gathered from the included studies was analysed through frequency counting of articles. Qualitative content analyses were used, and synthesised results were reported narratively. Study results were presented as narratives and on tables.

### Patient and public involvement

Patients and the public were not involved in the design, conduct, reporting or dissemination of this research.

## RESULTS

### Search results

The search yielded 1633 studies. After excluding duplicates, a total of 1355 studies proceeded to screening. Screening was conducted in two stages: 1) the title and abstract and 2) the full text screening, by AN and FC. After an independent review the differences between the two reviewers were later discussed. A total of 29 met the criteria for review. The results on the inclusion and exclusion are presented in a PRISMA diagram (Figure 1).

**Figure 1.**
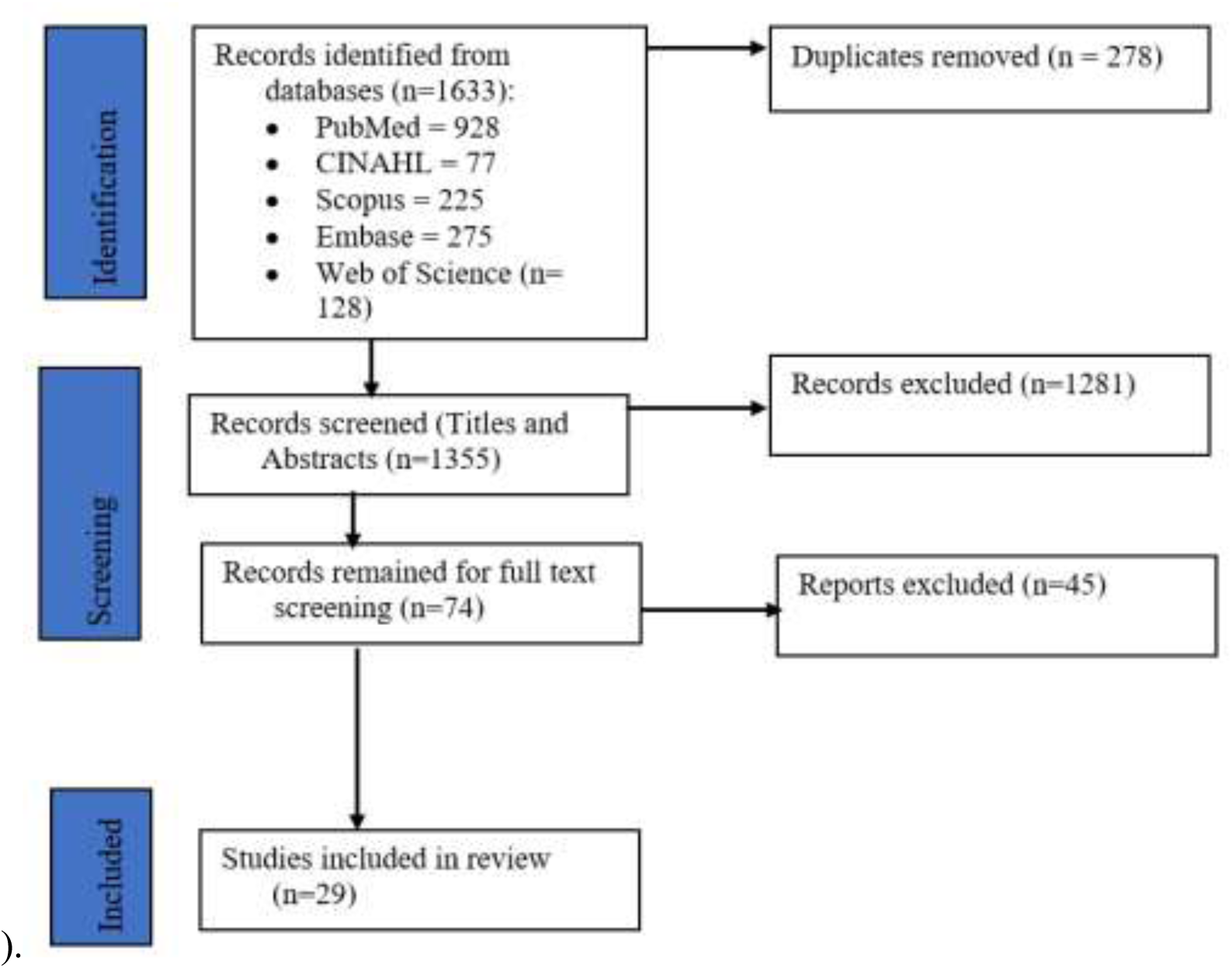
Preferred reporting items for systematic reviews and meta-analyses (PRISMA) flow chart.

### Critical appraisal

The critical appraisal was done between 26th May to 27th June 2024. Twenty-four studies were appraised as high, 6 as moderate and 3 as low. There was a high risk of bias for these studies. The detailed critical appraisal findings are presented in Table 1.

**Table 1:** Joanna Briggs Institute Critical Appraisal of Eligible Studies

| Cluster randomized control trial |  |  |  |  |  |  |  |  |  |  |  |  |  |  |
| --- | --- | --- | --- | --- | --- | --- | --- | --- | --- | --- | --- | --- | --- | --- |
| Author | Q1 | Q2 | Q3 | Q4 | Q5 | Q6 | Q7 | Q8 | Q9 | Q10 | Q11 | Q12 | Q13 |  |
| Aftab W et al 2021 | Y | U | N | Y | Y | Y | Y | Y | Y | Y | Y | Y | Y | High |
| Biemba et al 2020 | Y | Y | Y | Y | Y | Y | Y | Y | Y | Y | Y | Y | Y | High |
| Lazzerini.et al 2019 | Y | Y | Y | Y | Y | Y | Y | Y | Y | Y | Y | Y | Y | High |
| Rabani et al 2016 | Y | U | Y |  |  |  |  |  |  |  |  |  |  | Low |
| Bello 2013 | Y | Y | Y | Y | Y | Y | Y | Y | Y | Y | Y | Y | Y | High |
| Das et al 2015 | Y | Y | U | N | N | Y | U | Y | Y | Y | Y | Y | Y | Moderate |
| **Quasi experimental** |  |  |  |  |  |  |  |  |  |  |  |  |  |  |
| **Author** | **Q1** | **Q2** | **Q3** | **Q4** | **Q5** | **Q6** | **Q7** | **Q8** | **Q9** |  |  |  |  |  |
| Panda et al 2015 | Y | Y | Y | Y | Y | Y | Y | Y | Y |  |  |  |  | High |
| Adefolarin et al 2021 | Y | Y | Y | Y | Y | Y | Y | Y | Y |  |  |  |  | High |
| Usman et al 2023 | Y | Y | Y | Y | Y | Y | Y | Y | Y |  |  |  |  | High |
| Motaghi et al 2022 | Y | N | Y | N | Y | Y | Y | Y | Y |  |  |  |  | Moderate |
| Subramaniam et al 2018 | Y | N | N | Y | Y | Y | Y | Y | Y |  |  |  |  | Moderate |
| Aggarwal et al 2018 | Y | Y | Y | Y | Y | Y | Y | Y | Y |  |  |  |  | High |
| Eliades.et al 2019 | Y | N | Y | Y | Y | Y | Y | Y | Y |  |  |  |  | High |
| Meribe.et al 2020 | Y | Y | Y | Y | Y | Y | Y | Y | Y |  |  |  |  | High |
| Das. et al 2018 | Y | N | Y | Y | Y | Y | Y | Y | Y |  |  |  |  | High |
| Martin.et al 2019 | Y | Y | U | Y | Y | Y | Y | N | Y |  |  |  |  | Modérate |
| Mudogo.et al 2023 | Y | N | Y | Y | Y | Y | Y | Y | Y |  |  |  |  | High |
| Ameha et al 2015 | Y | N | N/A | Y | Y | Y | Y | Y | Y |  |  |  |  | Moderate |
| Worges et al 2018 | Y | N | N/A | Y | Y | Y | Y | Y | Y |  |  |  |  | Moderate |
| Renggli et al 2019 | Y | N | N/A | Y | Y | Y | Y | Y | Y |  |  |  |  | Moderate |
| Ashton et al 2023 | Y | N | N | U | Y | Y | Y | Y | Y |  |  |  |  | Low |
| **Qualitative study** |  |  |  |  |  |  |  |  |  |  |  |  |  |  |
| **Author** | **Q1** | **Q2** | **Q3** | **Q4** | **Q5** | **Q6** | **Q7** | **Q8** | **Q9** | **Q10** |  |  |  |  |
| Lutwana et al 2022 | Y | Y | Y | Y | Y | Y | Y | Y | Y | Y |  |  |  | High |
| Tanzil et al 2021 | Y | Y | Y | Y | Y | Y | Y | Y | Y | Y |  |  |  | High |
| Karuga et al 2019 | Y | Y | Y | Y | Y | Y | Y | Y | Y | Y |  |  |  | High |
| KoK et al 2018 | Y | Y | Y | Y | Y | Y | Y | Y | Y | Y |  |  |  | High |
| Mudogo et al 2023 | Y | Y | Y | Y | Y | Y | Y | Y | Y | Y |  |  |  | High |
| **Analytical Cross sectional studies** |  |  |  |  |  |  |  |  |  |  |  |  |  |  |
| **Author** | Q1 | Q2 | Q3 | Q4 | Q5 | Q6 | Q7 | Q8 |  |  |  |  |  |  |
| Mendhe et al 2019 | U | Y | Y | Y | Y | Y | Y | Y |  |  |  |  |  | High |
| Abdel -Razik 2021 | Y | Y | Y | Y | Y | Y | Y | Y |  |  |  |  |  | High |
| Rabbani.et al 2016 | Y | Y | Y | Y | NA | NA | Y | Y |  |  |  |  |  | High |
| Agoro. et al 2015 | Y | Y | Y | Y | NA | NA | Y | Y |  |  |  |  |  | High |
| Kok.et al 2018 | Y | Y | Y | Y | NA | NA | Y | Y |  |  |  |  |  | High |
| **Cohort studies** |  |  |  |  |  |  |  |  |  |  |  |  |  |  |
| **Author** | Q1 | Q2 | Q3 | Q4 | Q5 | Q6 | Q7 | Q8 | Q9 | Q10 | Q11 |  |  |  |
| Das et al 2018 | U | Y | Y | Y | Y | U | N | Y | Y | Y | N |  |  | Low |
| Gopalakrishnan | Y | Y | Y | Y | Y | Y | Y | Y | Y | Y | Y |  |  | High |
| Ashton et al 2022 | N | Y | Y | Y | Y | Y | Y | Y | Y | Y | Y |  |  | High |
| N, no; U, unclear; Y, yes; NA, not applicable. |  |  |  |  |  |  |  |  |  |  |  |  |  |  |

### Study characteristics

Data from the review of 29 peer-reviewed papers (8,10–37) were included in this systematic review. The reviewed studies had diverse methodologies used (Table 1): Five (8, 23, 24, 26, 31) of the studies were qualitative studies, six were cluster randomised control trials (12, 17–19, 25, 34), three were cohort studies (16, 20, 22), five were analytical cross-sectional studies (10, 14, 24, 28, 34), and fifteen were quasi-experimental studies (11, 13, 15, 16, 19, 21, 27, 29–33, 35–37). All 29 papers were conducted in PHC facilities in low- and middle-income countries (Figure 2). The papers had a mix of supportive supervision conducted in a variety of programmes as follows: 5 Malaria (17,19,21,27,37), 4 Immunisation (8,20,28,32), 5 PHC (10,24,26,30,33), 4 Integrated Community Case Management (ICCM) (12,15,16,18), 2 Nutrition (22,25), 1 Maternal Newborn and Child Health (MNCH) (34), 1 Mental Health (11), 1 Community Health Services (23), 1 IDSR (36), 1 IMNCI (13), 1 WASH (35), 1 TB (29), 1 Health Products and Technologies (31) and 1 Medicine Management (14) (Table 4).

### Country distribution of identified studies

The review of the studies identified that the studies were from the following: Egypt, Ethiopia, Pakistan, India, Kenya, Zambia, Nigeria, Mozambique, Tanzania and Malawi. Figure 2 shows the actual distribution of the studies.

**Figure 2:**
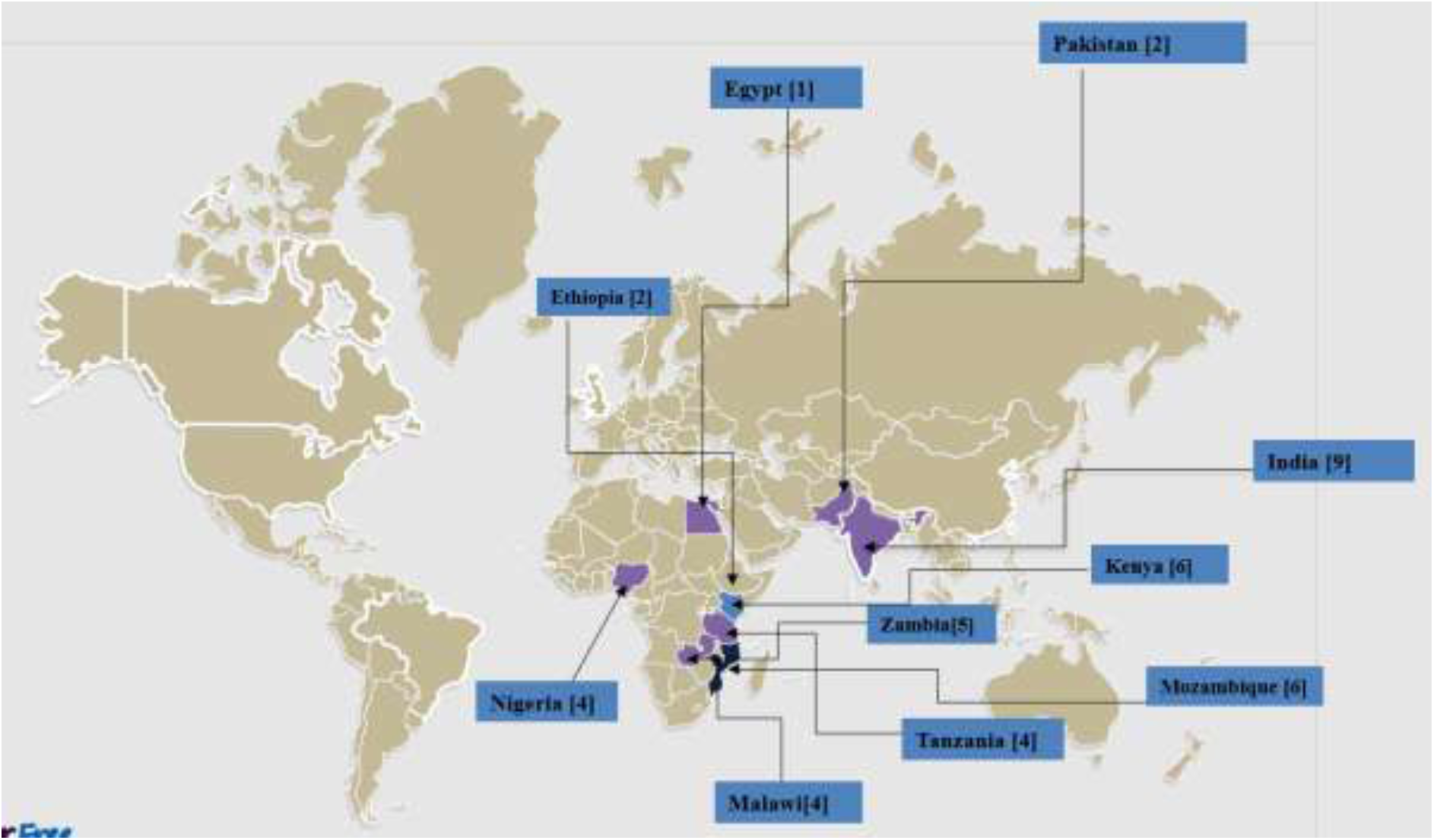
Distribution of Studies.

### Mechanism of Supportive supervision

Data from the review of 29 papers showed that there were 8 different approaches/mechanisms used when conducting SS in PHC in LMIC. The 29 papers were distributed in the 8 mechanisms as follows: 14% [n=4] used mechanism 1 (11, 17, 28, 1(11,17,28,35), 14% [n=4] used mechanism 2 (12, 26, 32, 36), 10.5% [n=3] of the papers used mechanism 3 (10, 19, 29), 24.5% [n=6] used mechanism 4 (8, 13, 13, 18, 22, 37), [n=5] used mechanism 5 (16, 23, 24, 27, 33), 7% [n=2] used mechanism 6 (14, 36), mechanism 7 (21, 30, 31), 10.5% [n=3] and mechanism 8 (15, 25) 3% [n=1]. Table 2 details the mechanisms for each paper.

**Table 2.**
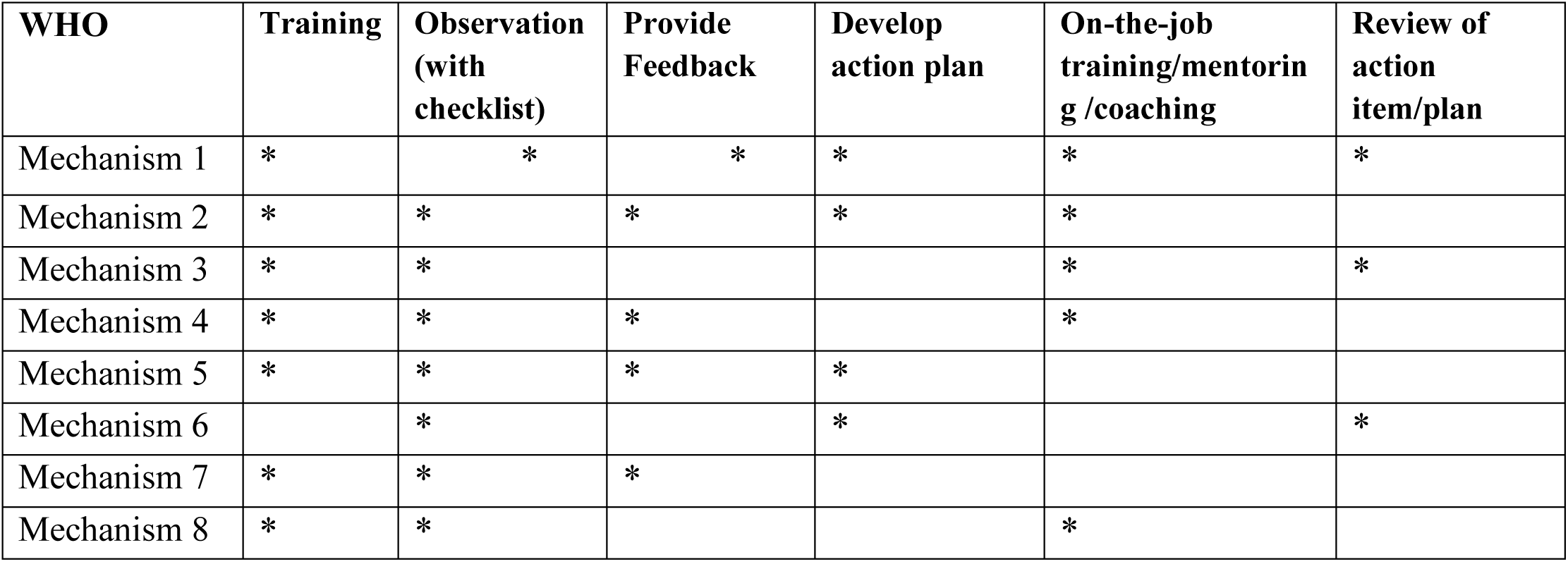
Mechanisms of supportive supervision.

| WHO | Training | Observation<br>(with<br>checklist) | Provide<br>Feedback | Develop<br>action plan | On-the-job<br>training/mentorin<br>g /coaching | Review of<br>action<br>item/plan |
| --- | --- | --- | --- | --- | --- | --- |
| Mechanism 1 | * | * | * | * | * | * |
| Mechanism 2 | * | * | * | * | * |  |
| Mechanism 3 | * | * |  |  | * | * |
| Mechanism 4 | * | * | * |  | * |  |
| Mechanism 5 | * | * | * | * |  |  |
| Mechanism 6 |  | * |  | * |  | * |
| Mechanism 7 | * | * | * |  |  |  |
| Mechanism 8 | * | * |  |  | * |  |

### Effectiveness of mechanisms of supportive supervision

The majority of papers reviewed (69% [n=20]) used a mechanism used for SS that was effective; 3.5% [n=1] of the results were ineffective, while 28% [n=8] of the study results were inconclusive (Table 2). Results on the effectiveness of mechanisms of SS per mechanism showed that in Mechanism 1, 10.5% [n=3] were effective while 3.5% [n=1] were inconclusive. Mechanism 2 results showed that 7% [n=2] of the papers had effective results, and 7% [n=2] were inconclusive. Mechanism 3 results showed 7% [n=2] were effective while 3.5% [n=1] were inconclusive. Mechanism 4 results showed 10.5% [n=3] were effective while 10.5% [n=1] were inconclusive. Mechanism 5 results showed 14% [n=4] were effective while 3.5% [n=1]. For Mechanism 6 results, 3.5% [n=1] were ineffective and 3.5% [n=1] were effective. Mechanism 7 results showed 10.5% [n=3] were effective, and so were mechanisms 8 results showed that 7% [n=2] (Table 3).

**Table 3:** Effectiveness of mechanism of supportive supervision.

| Mechanism | Ineffective | Effective | Inconclusive |
| --- | --- | --- | --- |
| 1 |  | (n=3) 10.5% (11,17,35) | [n=1] 3.5% (28) |
| 2 |  | (n=2) 7% (12,34) | [n=2] 7% (26,32) |
| 3 |  | (n=2) 7% (10,19) | [n=1] 3.5% (29) |
| 4 |  | (n=3) 10.5% (20,22,37) | [n=3] 10.5% (8,13,18) |
| 5 |  | (n=4) 14% (16,23,27,33) | [n=1] 3.5% (24) |
| 6 | [n=1] 3.5%(14) | (n=1) 3.5% (36) |  |
| 7 |  | (n=3) 10.5% (21,30,31) |  |
| 8 |  | (n=2) 7% (15,25) |  |
| TOTAL | 3.5% [n=1] | 69% [n=20] | 28% [n=8] |

### The role of supportive supervision in primary care performance

There are varying roles that SS plays in primary care performance (Table 4). This included improved knowledge, skills and practice in the different domains of programmes and improved health outcomes from service provision and treatment-seeking habits. It also was unable to confirm other positive roles of SS.

**Table 4:**
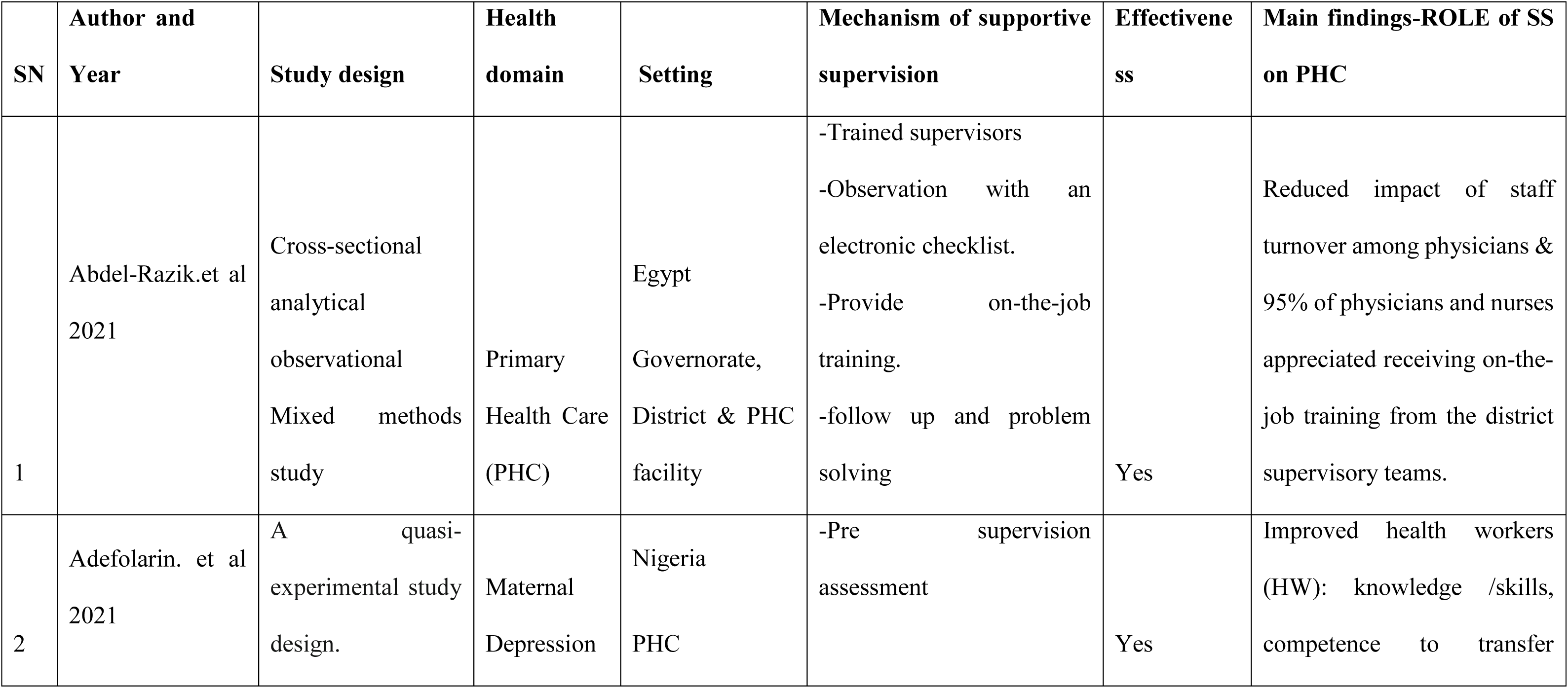
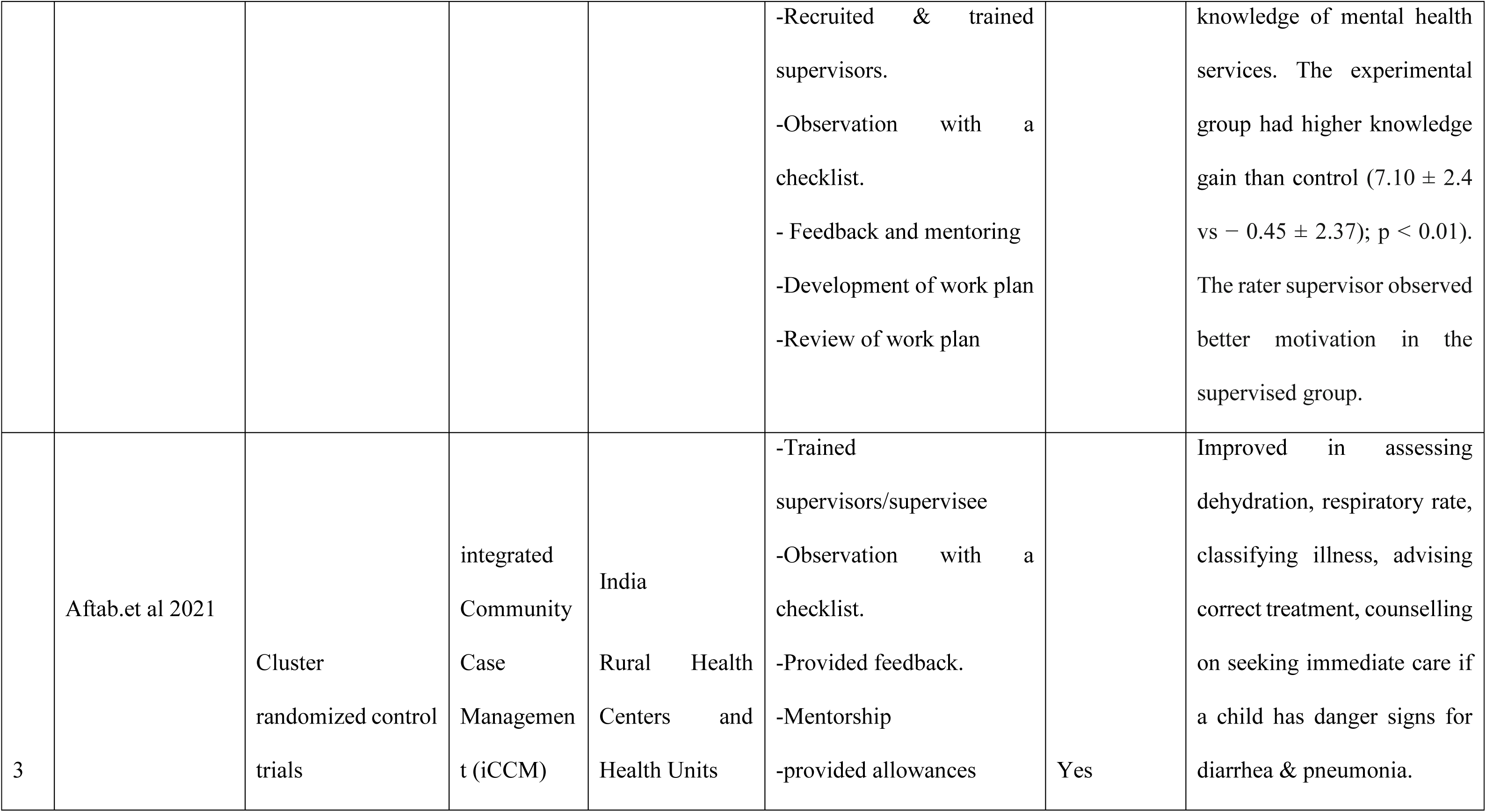
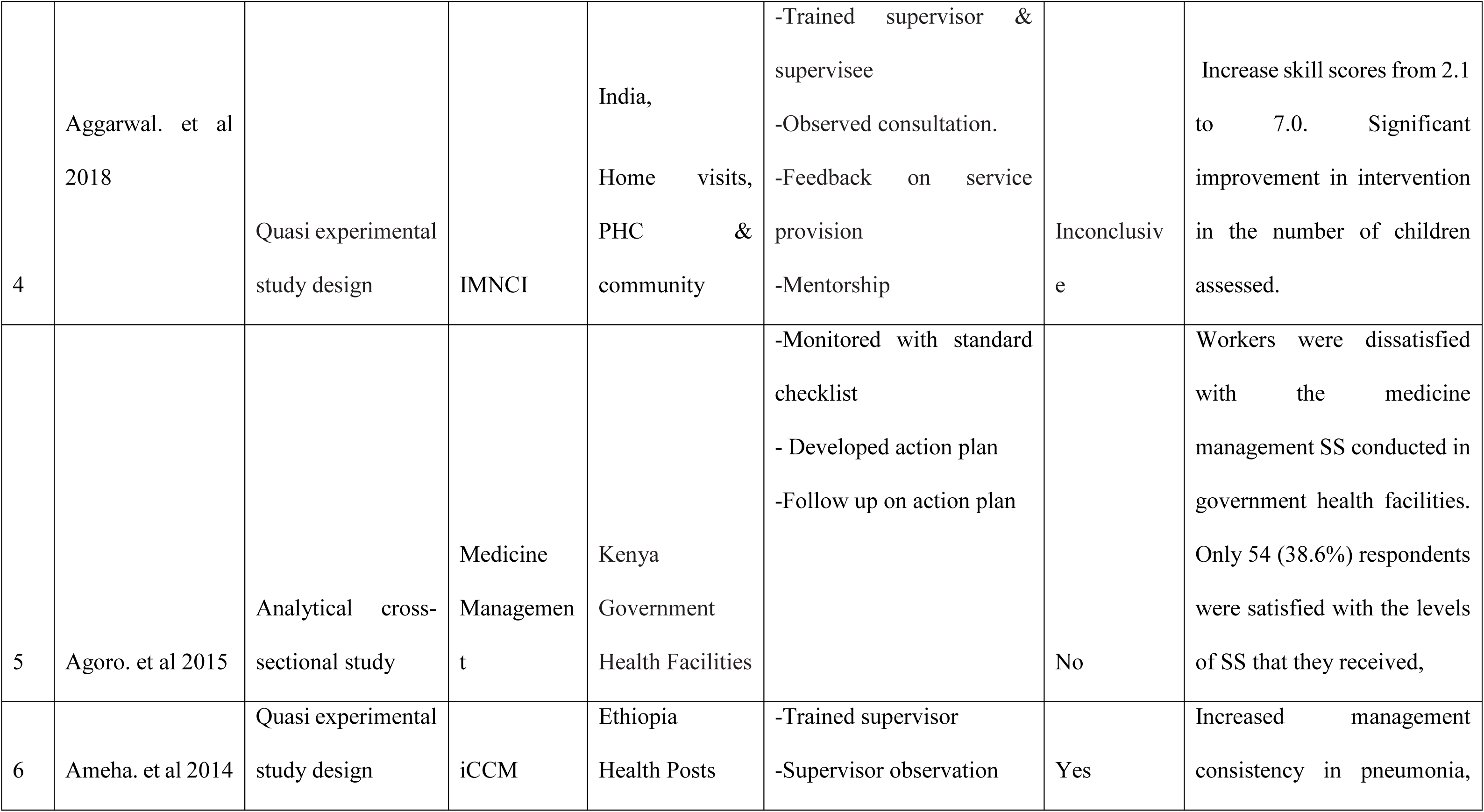
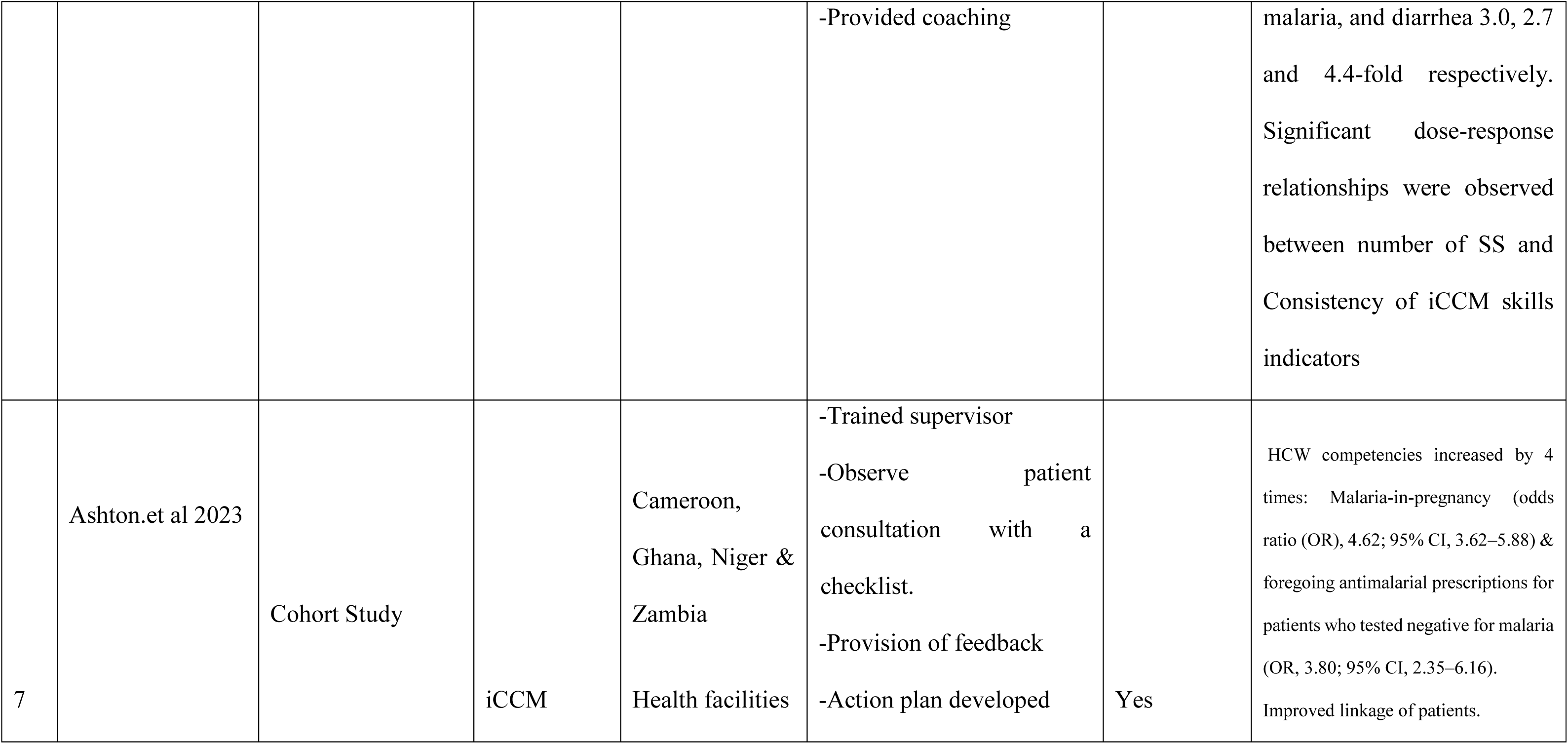
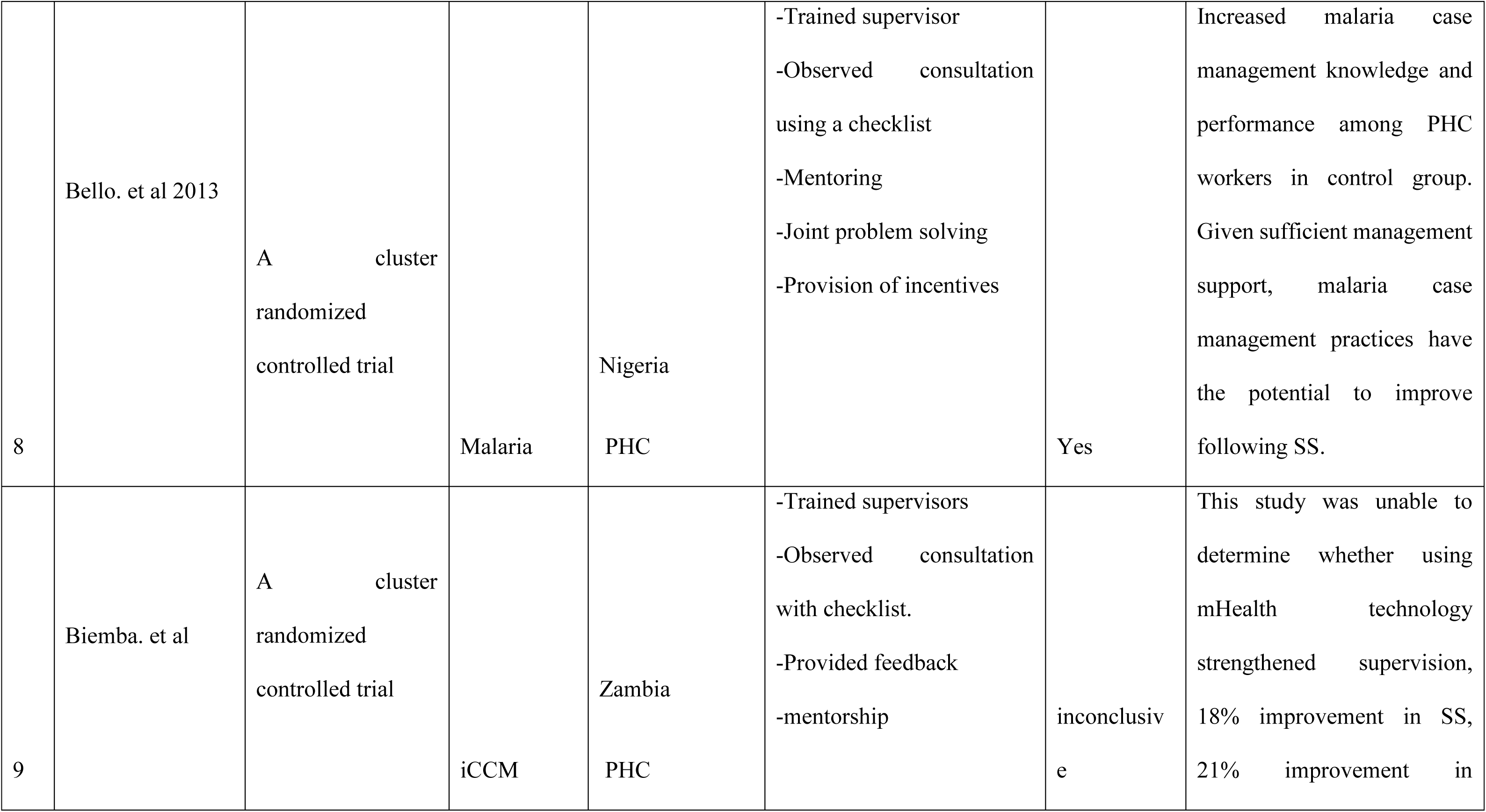
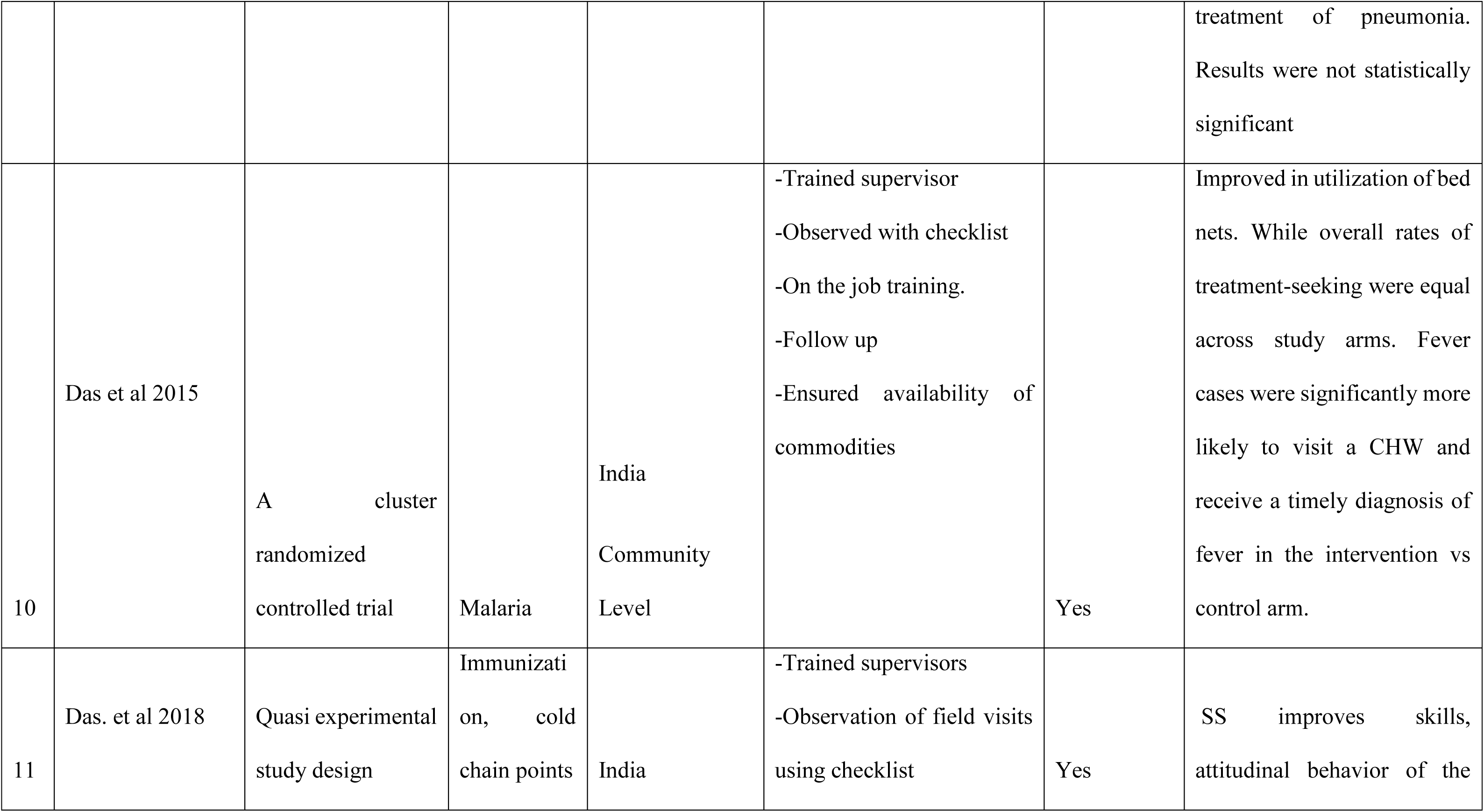
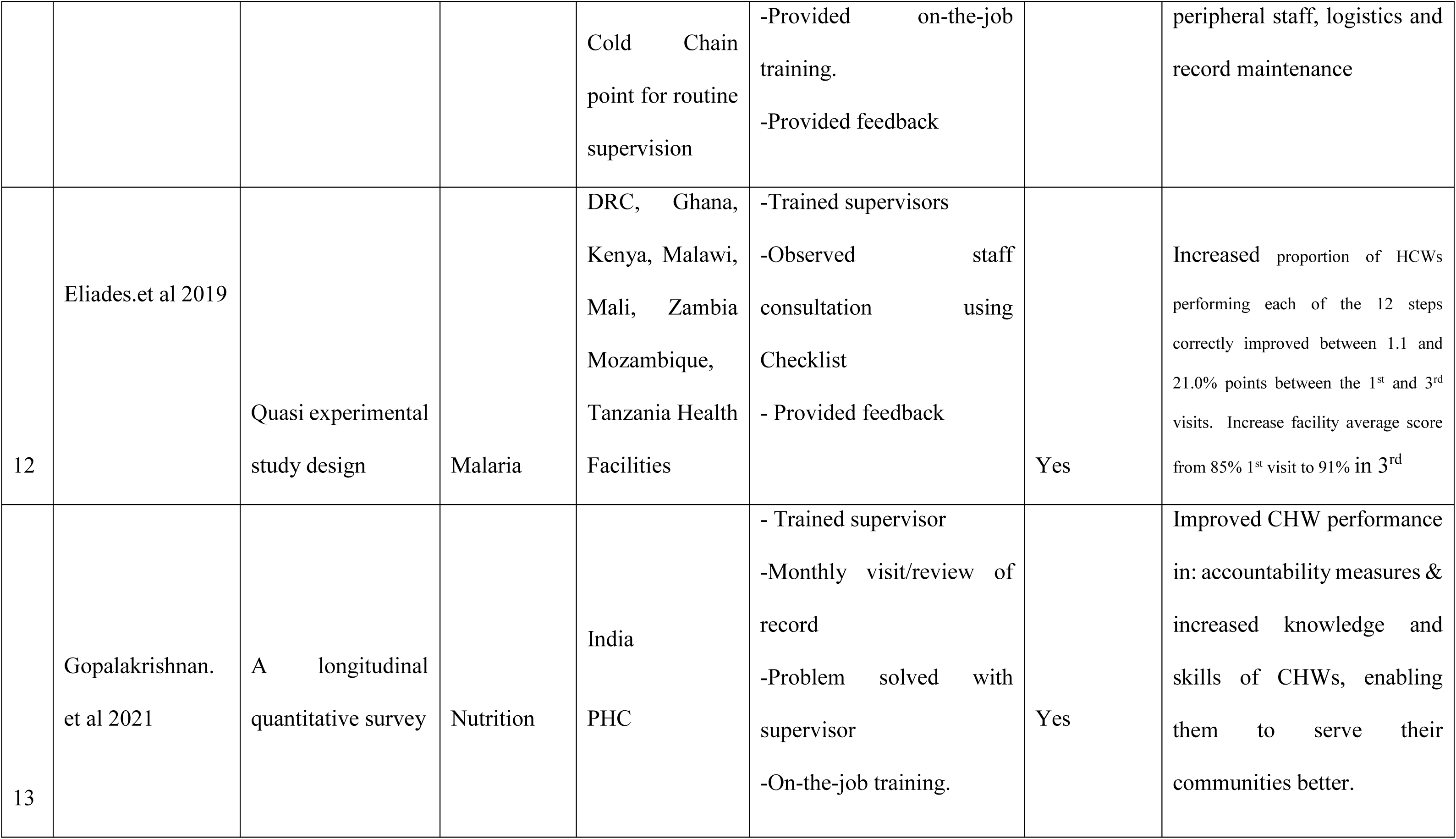
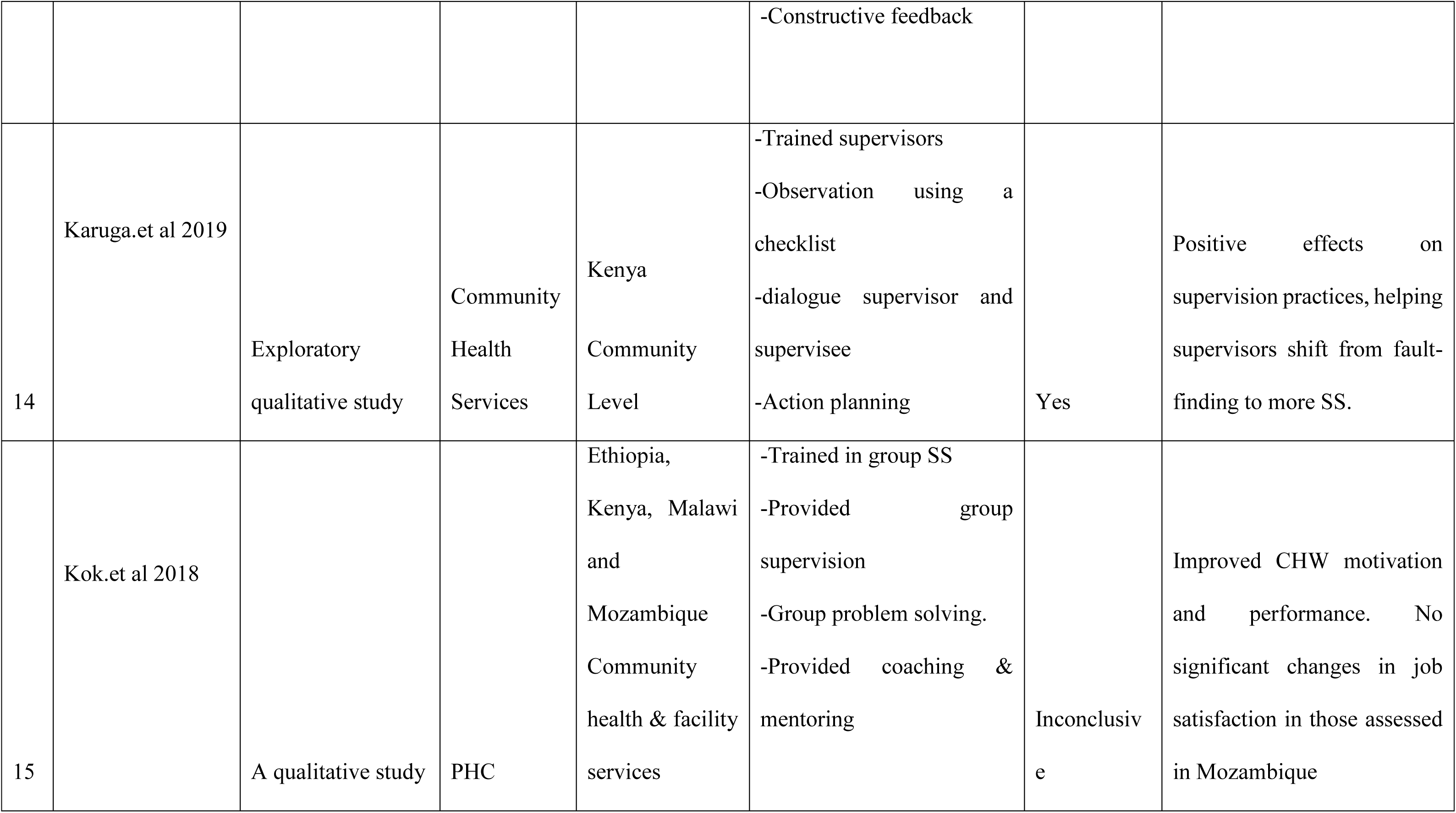
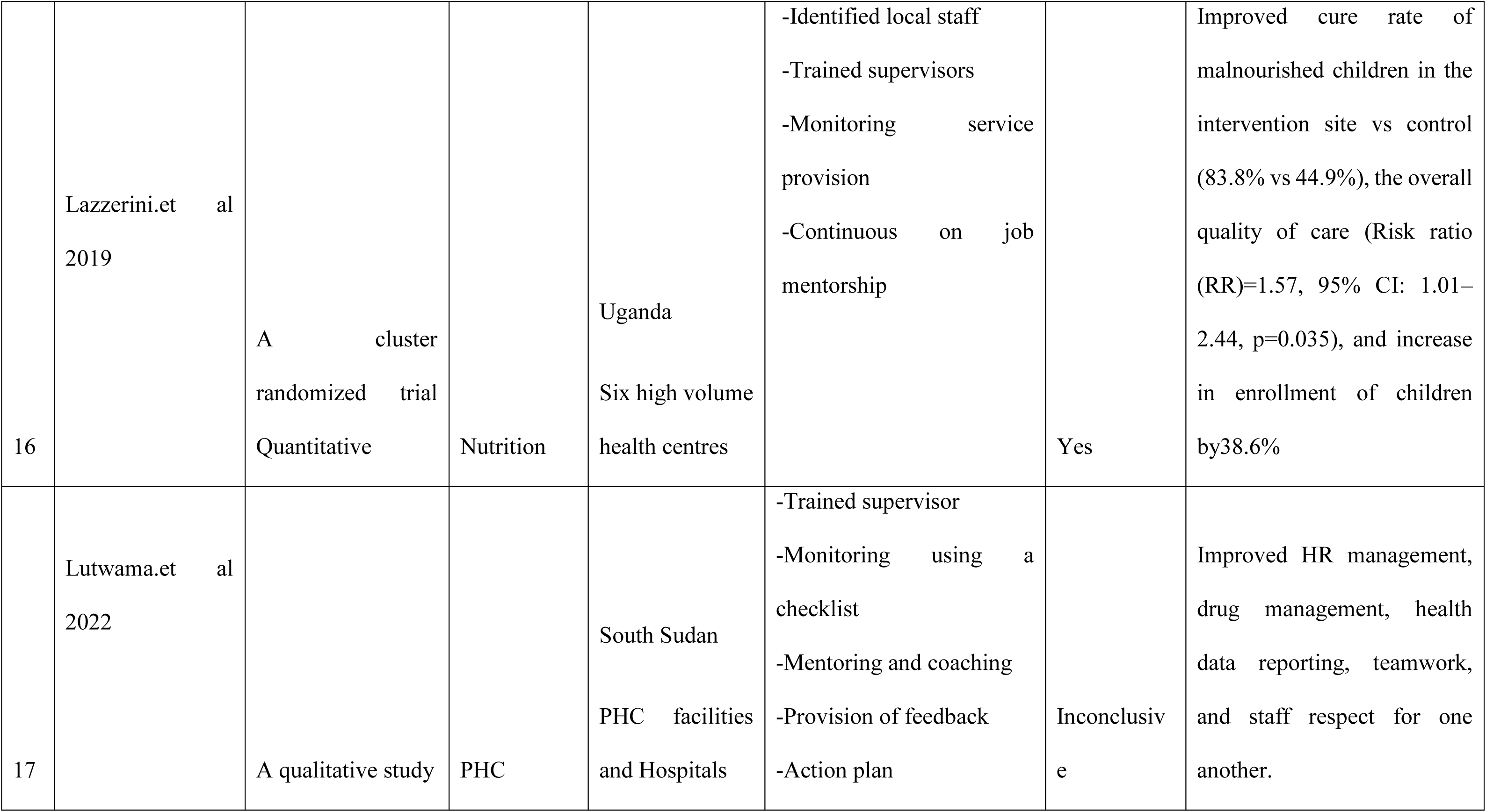
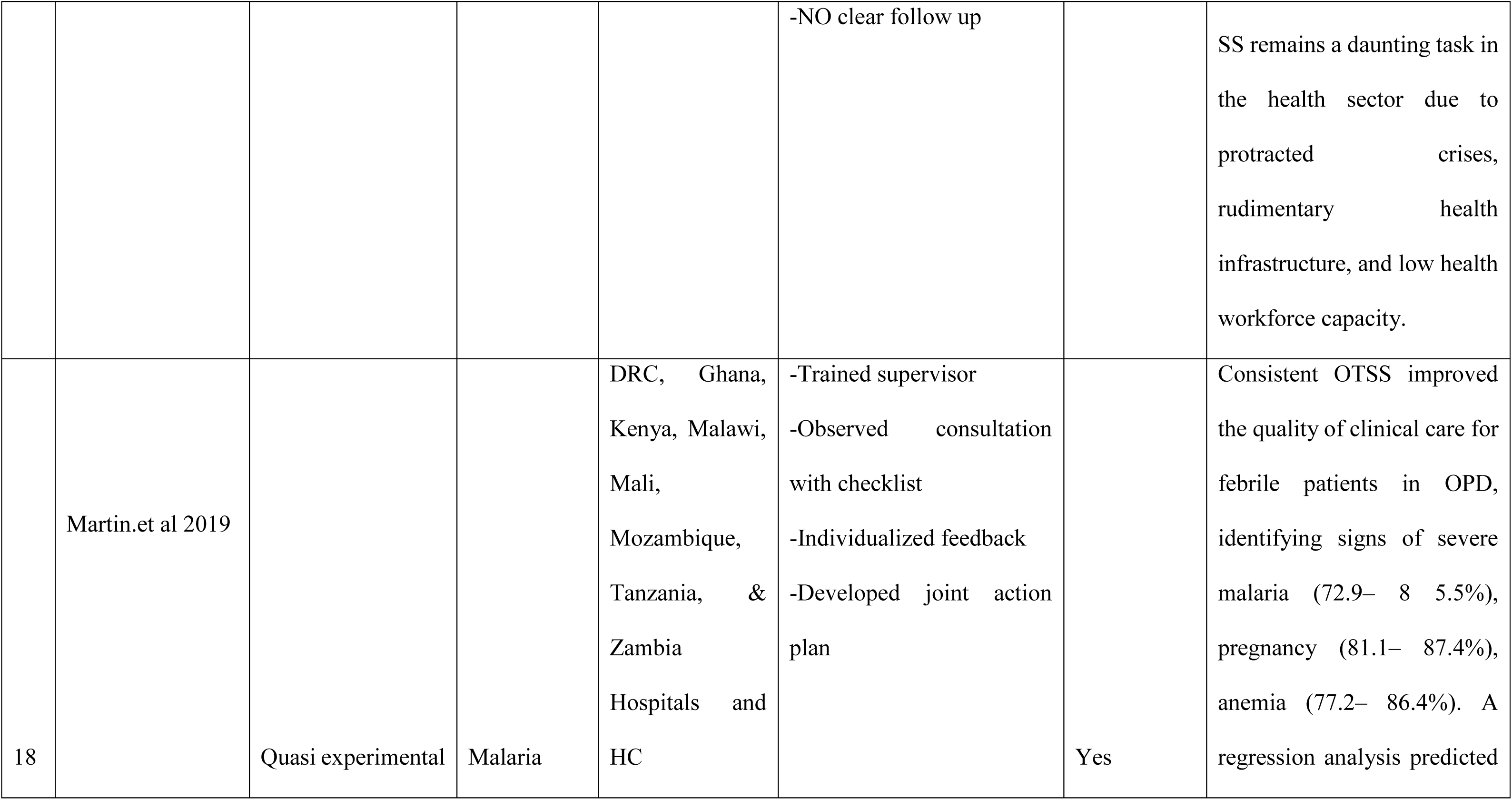
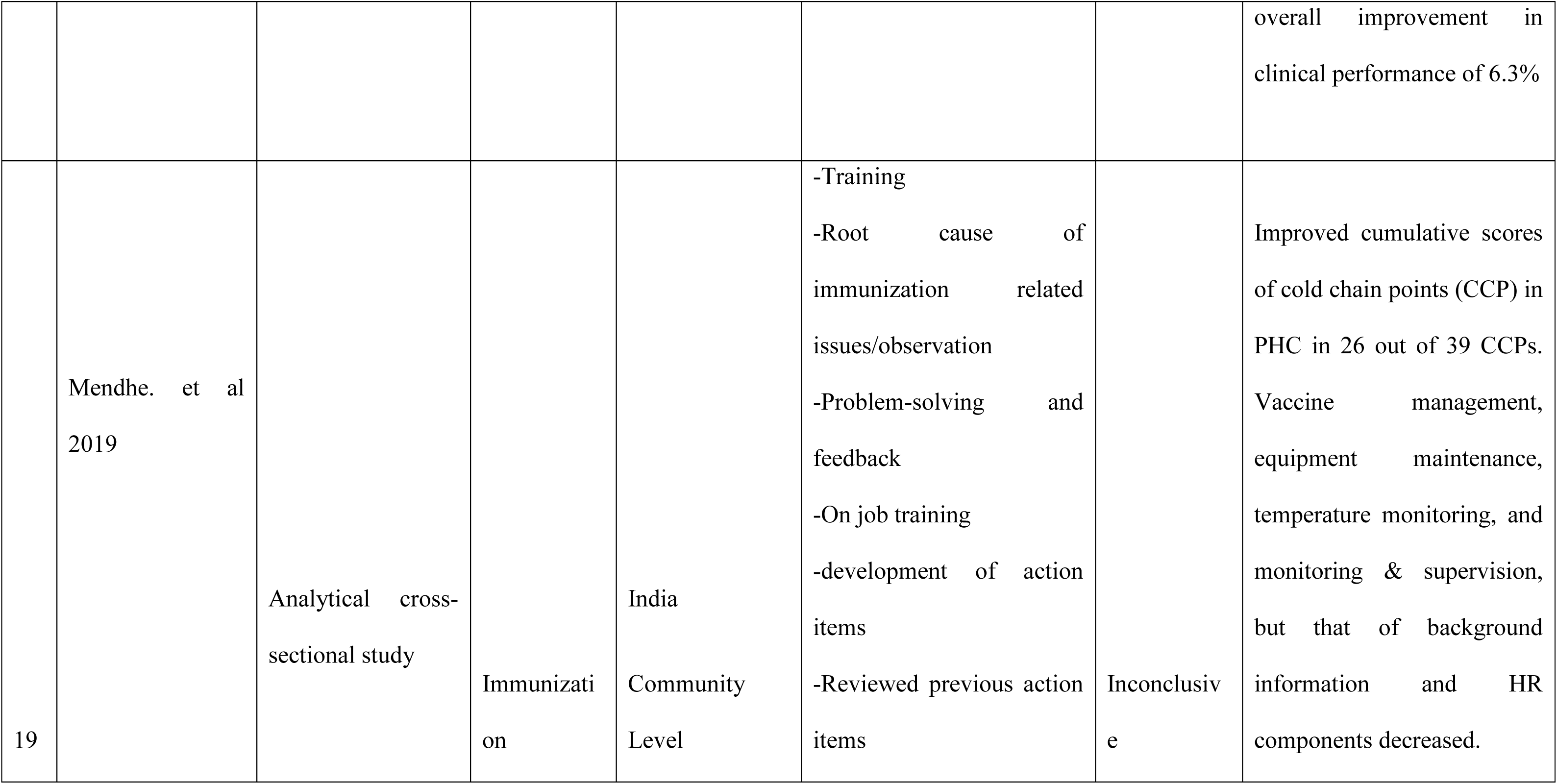
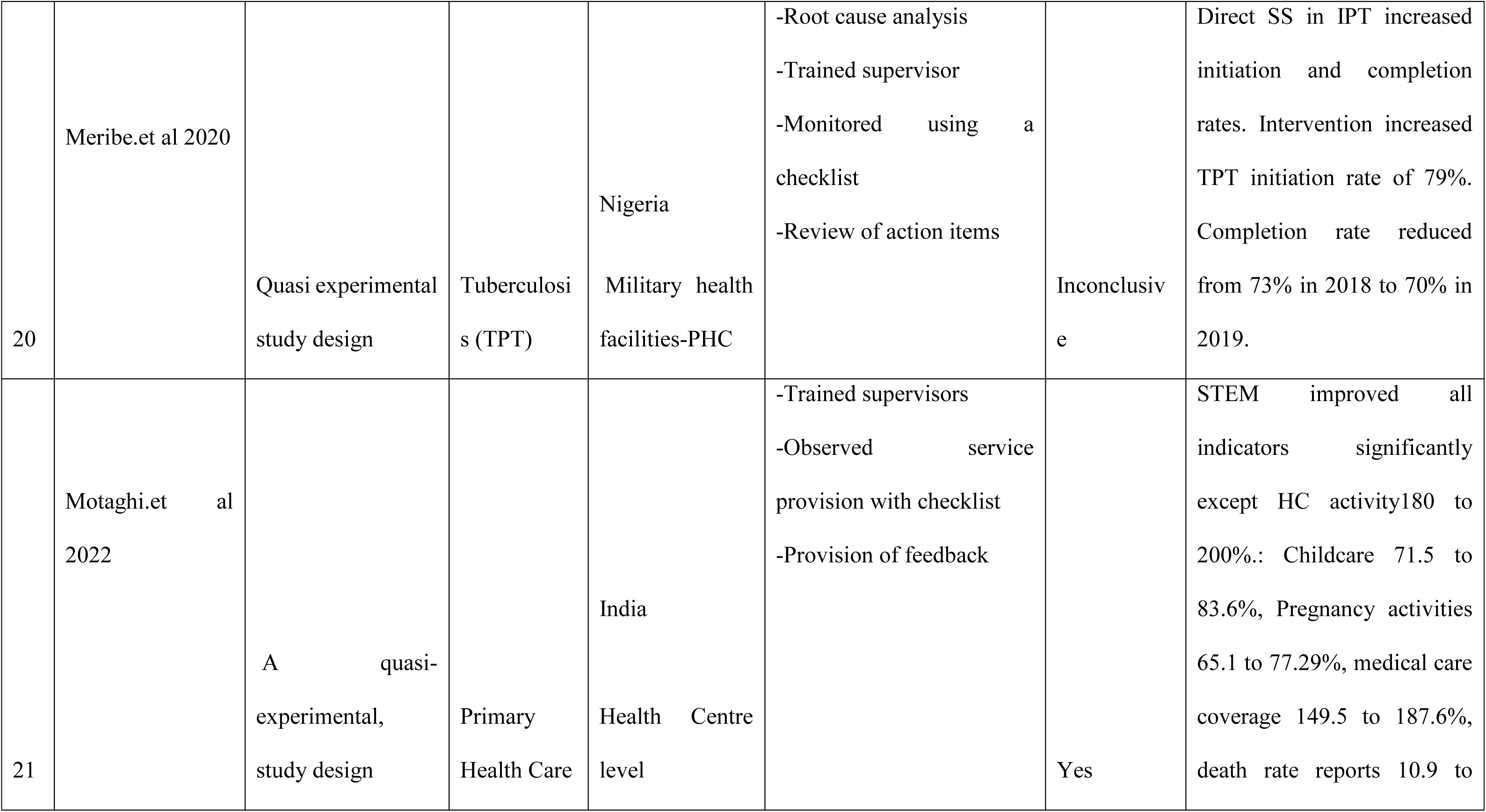
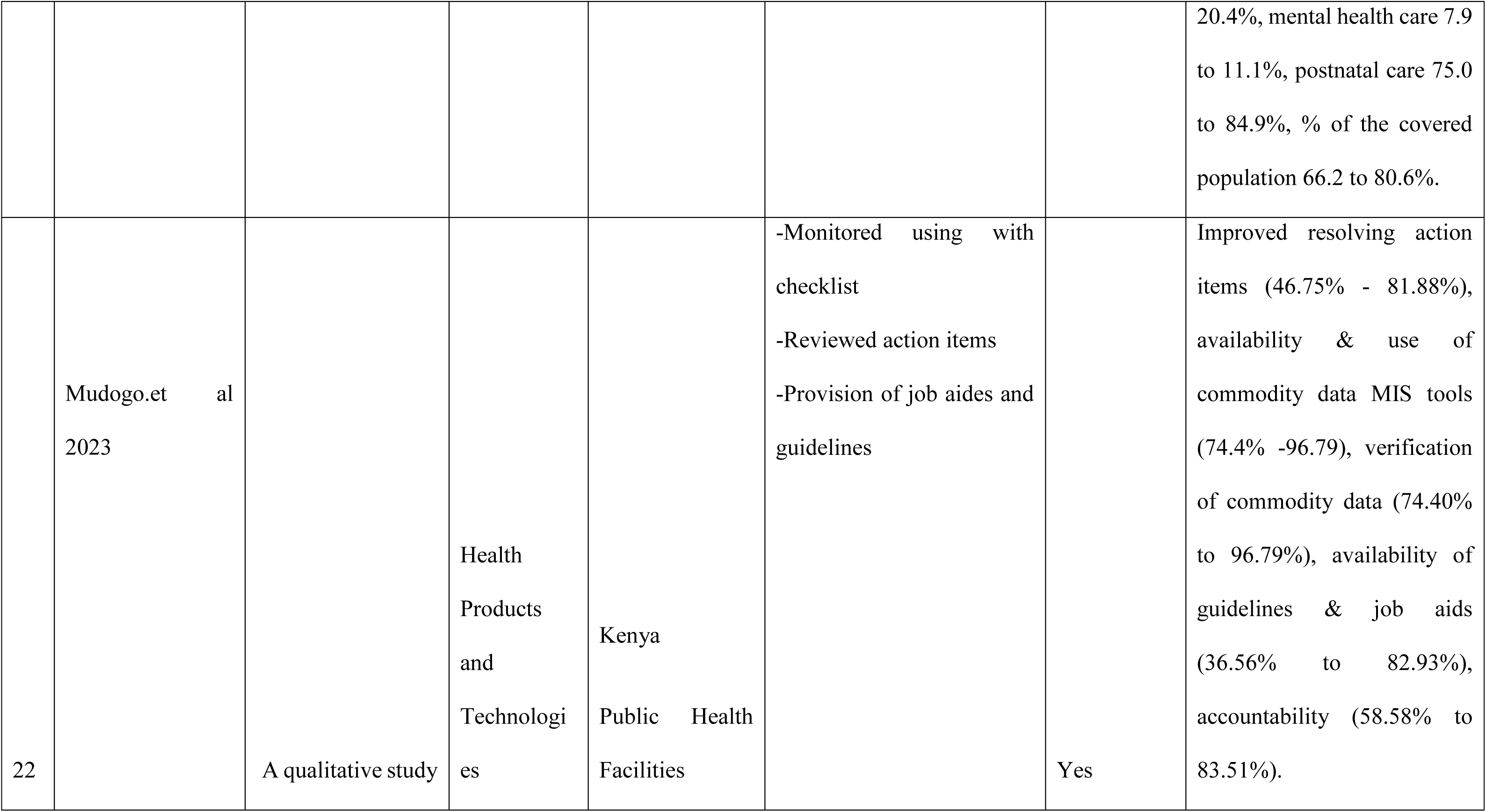
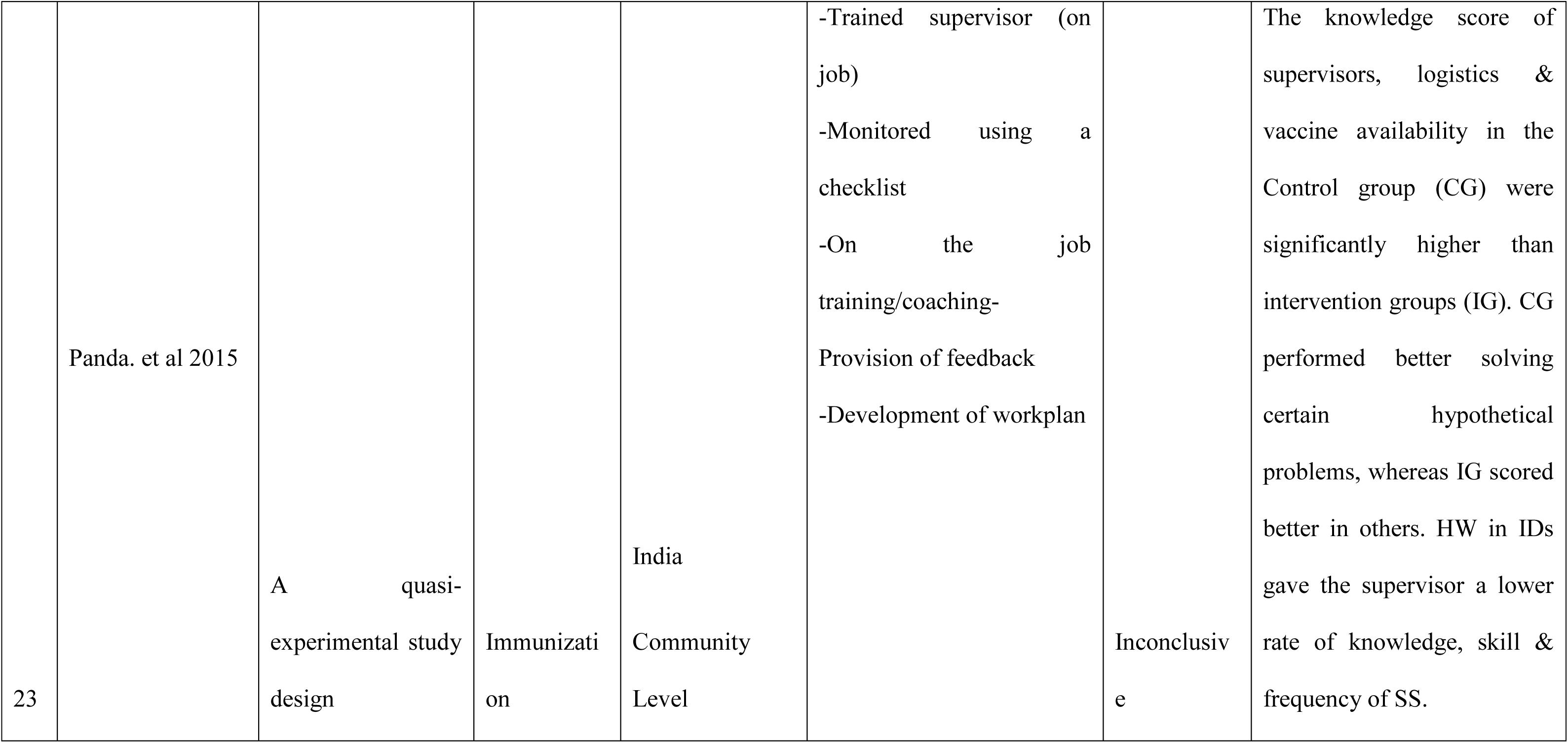
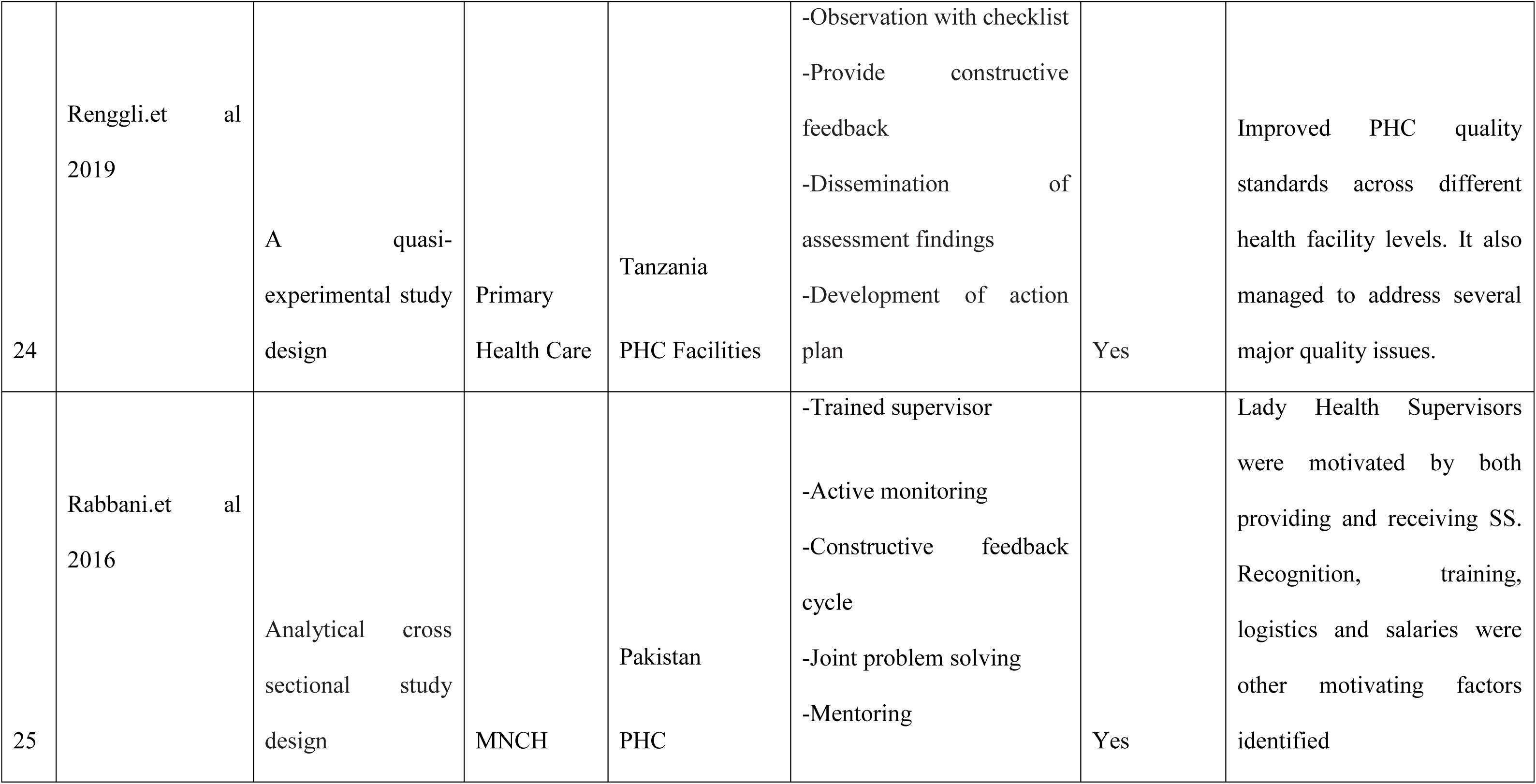
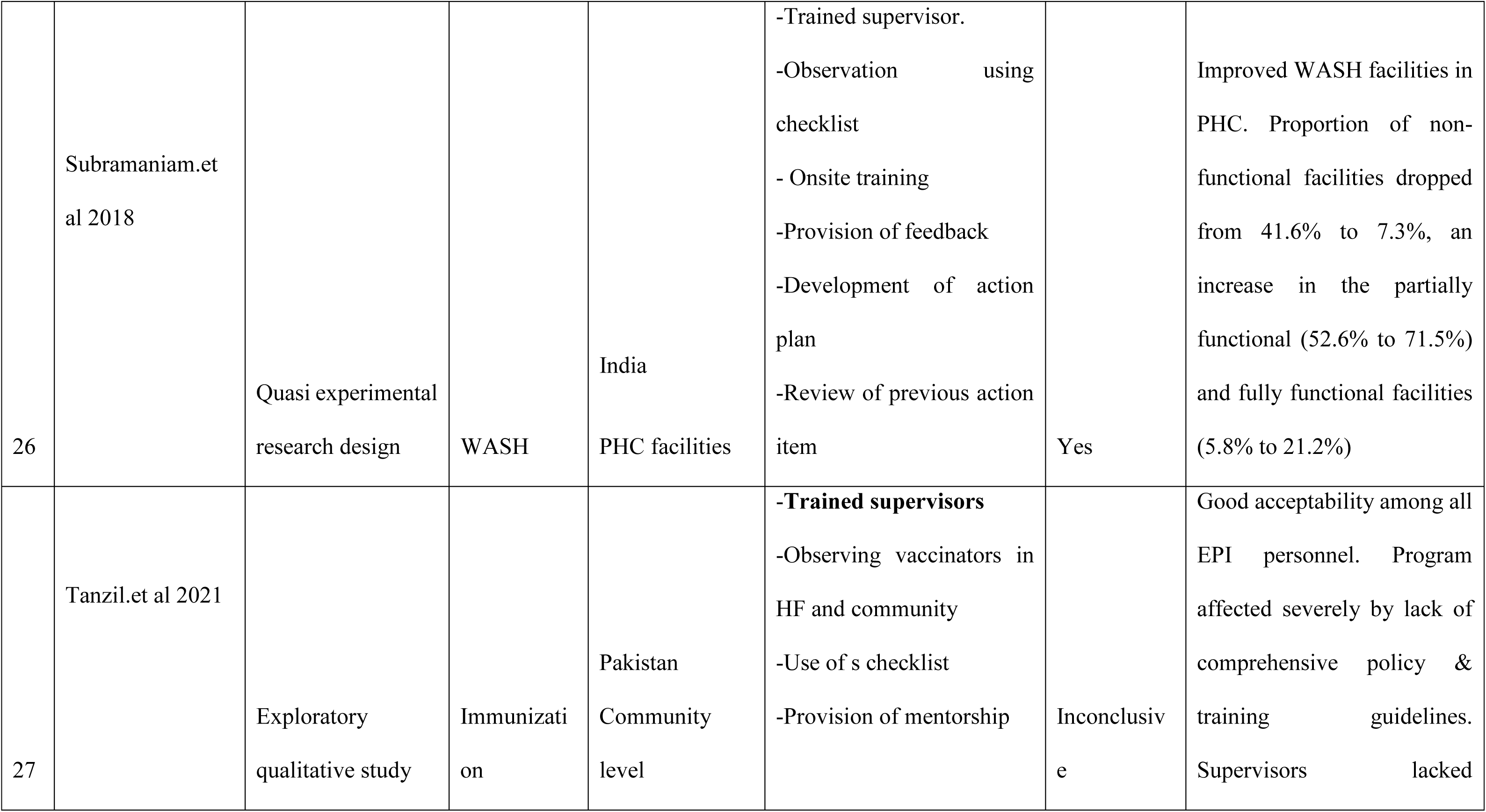
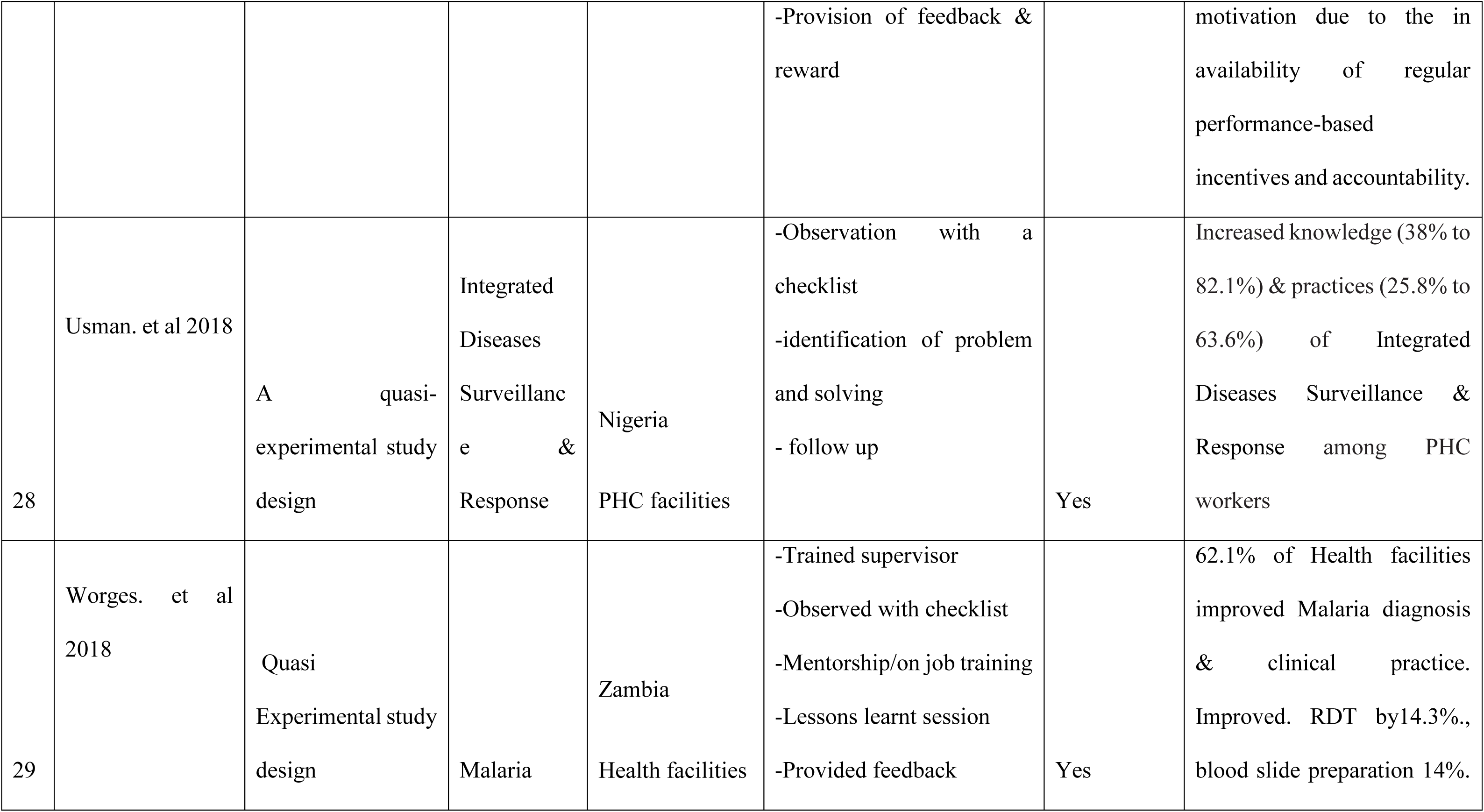
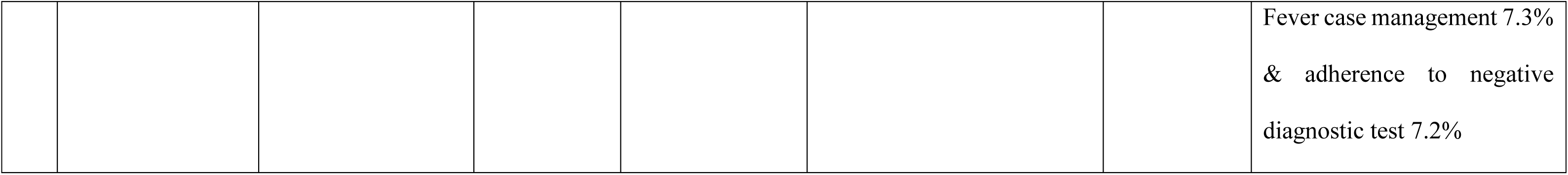
Role of supportive supervision in PHC in LMIC.

| SN | Author and Year | Study design | Health domain | Setting | Mechanism of supportive supervision | Effectiveness | Main findings-ROLE of SS on PHC |
| --- | --- | --- | --- | --- | --- | --- | --- |
| 1 | Abdel-Razik.et al<br>2021 | Cross-sectional analytical observational<br>Mixed methods study | Primary Health Care (PHC) | Egypt<br>Governorate, District & PHC facility | -Trained supervisors<br>-Observation with an electronic checklist.<br>-Provide on-the-job training.<br>-follow up and problem solving | Yes | Reduced impact of staff turnover among physicians & 95% of physicians and nurses appreciated receiving on-the-job training from the district supervisory teams. |
| 2 | Adefolarin. et al<br>2021 | A quasi-experimental study design. | Maternal Depression | Nigeria<br>PHC | -Pre supervision assessment | Yes | Improved health workers (HW): knowledge /skills, competence to transfer |
|  |  |  |  |  | -Recruited & trained supervisors.<br>-Observation with a checklist.<br>- Feedback and mentoring<br>-Development of work plan<br>-Review of work plan |  | knowledge of mental health services. The experimental group had higher knowledge gain than control ($7.10 \pm 2.4$ vs $-0.45 \pm 2.37$); $p < 0.01$).<br>The rater supervisor observed better motivation in the supervised group. |
| 3 | Aftab.et al 2021 | Cluster randomized control trials | integrated Community Case Management (iCCM) | India<br>Rural Health Centers and Health Units | -Trained supervisors/supervisee<br>-Observation with a checklist.<br>-Provided feedback.<br>-Mentorship<br>-provided allowances | Yes | Improved in assessing dehydration, respiratory rate, classifying illness, advising correct treatment, counselling on seeking immediate care if a child has danger signs for diarrhea & pneumonia. |
| 4 | Aggarwal. et al<br>2018 | Quasi experimental<br>study design | IMNCI | India,<br>Home visits,<br>PHC &<br>community | -Trained supervisor &<br>supervisee<br>-Observed consultation.<br>-Feedback on service<br>provision<br>-Mentorship | Inconclusiv<br>e | Increase skill scores from 2.1<br>to 7.0. Significant<br>improvement in intervention<br>in the number of children<br>assessed. |
| 5 | Agoro. et al 2015 | Analytical cross-sectional study | Medicine<br>Managemen<br>t | Kenya<br>Government<br>Health Facilities | -Monitored with standard<br>checklist<br>- Developed action plan<br>-Follow up on action plan | No | Workers were dissatisfied<br>with the medicine<br>management SS conducted in<br>government health facilities.<br>Only 54 (38.6%) respondents<br>were satisfied with the levels<br>of SS that they received, |
| 6 | Ameha. et al 2014 | Quasi experimental<br>study design | iCCM | Ethiopia<br>Health Posts | -Trained supervisor<br>-Supervisor observation | Yes | Increased management<br>consistency in pneumonia, |
|  |  |  |  |  | -Provided coaching |  | malaria, and diarrhea 3.0, 2.7 and 4.4-fold respectively. Significant dose-response relationships were observed between number of SS and Consistency of iCCM skills indicators |
| 7 | Ashton.et al 2023 | Cohort Study | iCCM | Cameroon,<br>Ghana, Niger &<br>Zambia<br>Health facilities | -Trained supervisor<br>-Observe patient consultation with a checklist.<br>-Provision of feedback<br>-Action plan developed | Yes | HCW competencies increased by 4 times: Malaria-in-pregnancy (odds ratio (OR), 4.62; 95% CI, 3.62–5.88) & foregoing antimalarial prescriptions for patients who tested negative for malaria (OR, 3.80; 95% CI, 2.35–6.16).<br>Improved linkage of patients. |
| 8 | Bello. et al 2013 | A cluster randomized controlled trial | Malaria | Nigeria<br>PHC | -Trained supervisor<br>-Observed consultation using a checklist<br>-Mentoring<br>-Joint problem solving<br>-Provision of incentives | Yes | Increased malaria case management knowledge and performance among PHC workers in control group. Given sufficient management support, malaria case management practices have the potential to improve following SS. |
| 9 | Biemba. et al | A cluster randomized controlled trial | iCCM | Zambia<br>PHC | -Trained supervisors<br>-Observed consultation with checklist.<br>-Provided feedback<br>-mentorship | inconclusive | This study was unable to determine whether using mHealth technology strengthened supervision, 18% improvement in SS, 21% improvement in treatment of pneumonia.<br>Results were not statistically significant |
| 10 | Das et al 2015 | A cluster randomized controlled trial | Malaria | India<br>Community Level | -Trained supervisor<br>-Observed with checklist<br>-On the job training.<br>-Follow up<br>-Ensured availability of commodities | Yes | Improved in utilization of bed nets. While overall rates of treatment-seeking were equal across study arms. Fever cases were significantly more likely to visit a CHW and receive a timely diagnosis of fever in the intervention vs control arm. |
| 11 | Das. et al 2018 | Quasi experimental study design | Immunization, cold chain points | India | -Trained supervisors<br>-Observation of field visits using checklist | Yes | SS improves skills, attitudinal behavior of the |
|  |  |  |  | Cold Chain point for routine supervision | -Provided on-the-job training.<br>-Provided feedback |  | peripheral staff, logistics and record maintenance |
| 12 | Eliades.et al 2019 | Quasi experimental study design | Malaria | DRC, Ghana, Kenya, Malawi, Mali, Zambia Mozambique, Tanzania Health Facilities | -Trained supervisors<br>-Observed staff consultation using Checklist<br>- Provided feedback | Yes | Increased proportion of HCWs performing each of the 12 steps correctly improved between 1.1 and 21.0% points between the 1<sup>st</sup> and 3<sup>rd</sup> visits. Increase facility average score from 85% 1<sup>st</sup> visit to 91% in 3<sup>rd</sup> |
| 13 | Gopalakrishnan. et al 2021 | A longitudinal quantitative survey | Nutrition | India<br>PHC | - Trained supervisor<br>-Monthly visit/review of record<br>-Problem solved with supervisor<br>-On-the-job training. | Yes | Improved CHW performance in: accountability measures & increased knowledge and skills of CHWs, enabling them to serve their communities better. |
|  |  |  |  |  | -Constructive feedback |  |  |
| 14 | Karuga.et al 2019 | Exploratory qualitative study | Community Health Services | Kenya<br>Community Level | -Trained supervisors<br>-Observation using a checklist<br>-dialogue supervisor and supervisee<br>-Action planning | Yes | Positive effects on supervision practices, helping supervisors shift from fault-finding to more SS. |
| 15 | Kok.et al 2018 | A qualitative study | PHC | Ethiopia,<br>Kenya, Malawi and<br>Mozambique<br>Community health & facility services | -Trained in group SS<br>-Provided group supervision<br>-Group problem solving.<br>-Provided coaching & mentoring | Inconclusive | Improved CHW motivation and performance. No significant changes in job satisfaction in those assessed in Mozambique |
| 16 | Lazzerini.et al<br>2019 | A cluster<br>randomized trial<br>Quantitative | Nutrition | Uganda<br>Six high volume<br>health centres | -Identified local staff<br>-Trained supervisors<br>-Monitoring service provision<br>-Continuous on job mentorship | Yes | Improved cure rate of malnourished children in the intervention site vs control (83.8% vs 44.9%), the overall quality of care (Risk ratio (RR)=1.57, 95% CI: 1.01–2.44, p=0.035), and increase in enrollment of children by 38.6% |
| 17 | Lutwama.et al<br>2022 | A qualitative study | PHC | South Sudan<br>PHC facilities<br>and Hospitals | -Trained supervisor<br>-Monitoring using a checklist<br>-Mentoring and coaching<br>-Provision of feedback<br>-Action plan | Inconclusive | Improved HR management, drug management, health data reporting, teamwork, and staff respect for one another. |
|  |  |  |  |  | -NO clear follow up |  | SS remains a daunting task in the health sector due to protracted crises, rudimentary health infrastructure, and low health workforce capacity. |
| 18 | Martin.et al 2019 | Quasi experimental | Malaria | DRC, Ghana, Kenya, Malawi, Mali, Mozambique, Tanzania, & Zambia Hospitals and HC | -Trained supervisor<br>-Observed consultation with checklist<br>-Individualized feedback<br>-Developed joint action plan | Yes | Consistent OTSS improved the quality of clinical care for febrile patients in OPD, identifying signs of severe malaria (72.9– 85.5%), pregnancy (81.1– 87.4%), anemia (77.2– 86.4%). A regression analysis predicted overall improvement in clinical performance of 6.3% |
| 19 | Mendhe. et al<br>2019 | Analytical cross-sectional study | Immunization | India<br>Community Level | -Training<br>-Root cause of immunization related issues/observation<br>-Problem-solving and feedback<br>-On job training<br>-development of action items<br>-Reviewed previous action items | Inconclusive | Improved cumulative scores of cold chain points (CCP) in PHC in 26 out of 39 CCPs. Vaccine management, equipment maintenance, temperature monitoring, and monitoring & supervision, but that of background information and HR components decreased. |
| 20 | Meribe.et al 2020 | Quasi experimental study design | Tuberculosis (TPT) | Nigeria<br>Military health facilities-PHC | -Root cause analysis<br>-Trained supervisor<br>-Monitored using a checklist<br>-Review of action items | Inconclusive | Direct SS in IPT increased initiation and completion rates. Intervention increased TPT initiation rate of 79%. Completion rate reduced from 73% in 2018 to 70% in 2019. |
| 21 | Motaghi.et al 2022 | A quasi-experimental, study design | Primary Health Care | India<br>Health Centre level | -Trained supervisors<br>-Observed service provision with checklist<br>-Provision of feedback | Yes | STEM improved all indicators significantly except HC activity 180 to 200%.: Childcare 71.5 to 83.6%, Pregnancy activities 65.1 to 77.29%, medical care coverage 149.5 to 187.6%, death rate reports 10.9 to |
|  |  |  |  |  |  |  | 20.4%, mental health care 7.9 to 11.1%, postnatal care 75.0 to 84.9%, % of the covered population 66.2 to 80.6%. |
| 22 | Mudogo et al<br>2023 | A qualitative study | Health Products and Technologies | Kenya<br>Public Health Facilities | -Monitored using with checklist<br>-Reviewed action items<br>-Provision of job aides and guidelines | Yes | Improved resolving action items (46.75% - 81.88%), availability & use of commodity data MIS tools (74.4% -96.79), verification of commodity data (74.40% to 96.79%), availability of guidelines & job aids (36.56% to 82.93%), accountability (58.58% to 83.51%). |
| 23 | Panda. et al 2015 | A quasi-experimental study design | Immunization | India<br>Community Level | -Trained supervisor (on job)<br>-Monitored using a checklist<br>-On the job training/coaching-Provision of feedback<br>-Development of workplan | Inconclusive | The knowledge score of supervisors, logistics & vaccine availability in the Control group (CG) were significantly higher than intervention groups (IG). CG performed better solving certain hypothetical problems, whereas IG scored better in others. HW in IDs gave the supervisor a lower rate of knowledge, skill & frequency of SS. |
| 24 | Renggli.et al<br>2019 | A quasi-experimental study design | Primary Health Care | Tanzania<br>PHC Facilities | -Observation with checklist<br>-Provide constructive feedback<br>-Dissemination of assessment findings<br>-Development of action plan | Yes | Improved PHC quality standards across different health facility levels. It also managed to address several major quality issues. |
| 25 | Rabbani.et al<br>2016 | Analytical cross sectional study design | MNCH | Pakistan<br>PHC | -Trained supervisor<br>-Active monitoring<br>-Constructive feedback cycle<br>-Joint problem solving<br>-Mentoring | Yes | Lady Health Supervisors were motivated by both providing and receiving SS. Recognition, training, logistics and salaries were other motivating factors identified |
| 26 | Subramaniam.et al 2018 | Quasi experimental research design | WASH | India<br>PHC facilities | -Trained supervisor.<br>-Observation using checklist<br>- Onsite training<br>-Provision of feedback<br>-Development of action plan<br>-Review of previous action item | Yes | Improved WASH facilities in PHC. Proportion of non-functional facilities dropped from 41.6% to 7.3%, an increase in the partially functional (52.6% to 71.5%) and fully functional facilities (5.8% to 21.2%) |
| 27 | Tanzil.et al 2021 | Exploratory qualitative study | Immunization | Pakistan<br>Community level | -**Trained supervisors**<br>-Observing vaccinators in HF and community<br>-Use of s checklist<br>-Provision of mentorship | Inconclusive | Good acceptability among all EPI personnel. Program affected severely by lack of comprehensive policy & training guidelines. Supervisors lacked |
|  |  |  |  |  | -Provision of feedback & reward |  | motivation due to the in availability of regular performance-based incentives and accountability. |
| 28 | Usman. et al 2018 | A quasi-experimental study design | Integrated Diseases Surveillance & Response | Nigeria<br>PHC facilities | -Observation with a checklist<br>-identification of problem and solving<br>- follow up | Yes | Increased knowledge (38% to 82.1%) & practices (25.8% to 63.6%) of Integrated Diseases Surveillance & Response among PHC workers |
| 29 | Worges. et al 2018 | Quasi Experimental study design | Malaria | Zambia<br>Health facilities | -Trained supervisor<br>-Observed with checklist<br>-Mentorship/on job training<br>-Lessons learnt session<br>-Provided feedback | Yes | 62.1% of Health facilities improved Malaria diagnosis & clinical practice. Improved. RDT by 14.3%., blood slide preparation 14%. |
|  |  |  |  |  |  |  | Fever case management 7.3%<br>& adherence to negative<br>diagnostic test 7.2% |

## DISCUSSION

This systematic review aimed to examine the various mechanisms of supportive supervision (SS) employed in primary care settings, assess their effectiveness, and explore the role SS plays in enhancing primary care performance. The findings revealed that 1) multiple mechanisms are used to conduct SS; 2) while many of these mechanisms were effective, some produced ineffective and inconclusive outcomes; and 3) SS contributed to improvements in attitudinal behaviour, knowledge, skills and practice, as well as in health and patient outcomes.

### 1. Mechanism of supportive supervision

The review analysed 29 studies employing quantitative, qualitative and mixed methods (Table 1) to examine the mechanisms of SS used in PHC in LMIC. Eight different approaches to conducting SS were identified. Despite some variations, the mechanisms mostly followed a similar pattern structure: PHC supervisors were external to the facility being assessed, and almost always the supervisors are affiliated with a higher-level institution such as district, regional or national level. Both supervisors and, in some cases, supervisees received training on how to conduct or engage in SS. Observation or monitoring was commonly guided by a checklist or standardised paper-based or electronic.

Variation in SS mechanisms is primarily observed in the steps following the observation or monitoring phase. In certain models, supervision ended after feedback was provided to supervisees. In others, supervisors provided additional support through mentorship, coaching or on-the-job training – although these activities were sometimes described without explicitly referencing the provision of feedback. In a third approach, supervision extended further to include joint development of action plans between supervisors and supervisees.

To assess alignment with the best practice, this review applied the World Health Organization’s (WHO) 2008 module of SS as a benchmark/gold standard. The WHO module looks at four core components for setting up an SS programme:1) Setting up an SS system 2) planning regular SS visits 3) conducting a supervisory visit and 4) following up on activities. This review focused on the third and fourth steps as they pertain directly to how SS is approached. From these two steps, six components are considered as essential to effective SS:

i. Training the supervisor on how to conduct supervision
ii. Use of checklist by Supervisors to observe and monitoring supervisee or facility performance
iii. Provision of feedback to the supervisees
iv. On the job training, mentoring, or coaching in the areas were performance gaps are identified.
v. Joint development of an action plan between the supervisor and supervisee to address observed gaps
vi. Review of previous action items, typically done at the beginning of the next supervision session.

Using these six components as a framework, the review compared the eight mechanisms identified against the WHO’s guidance (Table 1). The analysis revealed inconsistencies in how SS is implemented across contexts, suggesting that the WHO framework is not uniformly applied. This divergence highlights a need for greater standardisation in operationalising supportive supervision in PHC systems in LMICs.

### 2. Effectiveness of Supportive Supervision in Primary Health Care

The review of SS effectiveness identified three categories of outcomes: effective, ineffective or inconclusive. Of the 29 studies reviewed, 69% [n=20] reported the SS mechanism as effective, 28% [n=8] as ineffective and 3.5% [n=1] as inconclusive. The review categorised the studies based on SS impact on three domains: healthcare provider performance, health system strengthening and patient outcomes.

Mechanisms 1, 4, 5 and 7 were the most effective SS approaches, with Mechanism 5 showing the highest number (14%, n=4) of effective studies (Table 2). Notably, Mechanism 5 incorporated four of the six WHO components: training, observation, feedback and work plan development. Although it lacked two components, its effectiveness suggests that these four may be particularly influential. Interestingly, mechanism 1, which included **all six WHO components**, was also among the most effective, indicating that comprehensive approaches tend to yield better outcomes. However, the review also noted **variability in implementation**, with differences in how SS was conducted across mechanisms.

A key insight from the review was the **interchangeability** of certain components— particularly **feedback (component 3)** and **on-the-job training/mentoring (component 5)**. Feedback identifies areas for improvement, while mentoring addresses them directly. This suggests that these steps may be **functionally complementary** and not always required as separate actions.

Importantly, the review found that **effectiveness is not solely determined by the SS mechanism used**. For example, Mechanisms 1 through 5 showed both effective and inconclusive results, while Mechanism 6 had both effective and ineffective outcomes. This underscores the influence of **contextual factors**, such as time allocated for SS, consistency of supervision, availability of incentives (26, 21), and adequate human resources and medical supplies. However, one study (14) highlighted that **irregular supervisory visits**, **lack of standardised checklists**, and **poor follow-up on action items** contributed to poor outcomes. These are significant **barriers** to achieving positive results through SS.

In line with Donabedian’s framework, achieving quality health outcomes requires both adequate inputs and appropriate processes. The findings reinforce the inconsistencies in what constitutes an effective SS process and suggest that contextual and implementation factors are critical to success.

### 3. The role of supportive supervision on primary care performance

This systematic review examined the effectiveness of SS in enhancing primary care performance. The evidence from the reviewed studies highlights three key areas where SS contributes positively. This includes 1) improvement in health workers’ attitudinal behaviour, 2) enhancement of health workers’ knowledge and skills, and 3) improvement in patient outcome. Nonetheless, a few studies showed how SS lacked the ability to change the health workers’ behaviour or facility and patient health outcomes.

#### Improvement in health workers attitudinal behaviour

Supportive supervision was found to positively influence health workers’ attitudes, including motivation and job satisfaction (24,34), interpersonal respect among staff (20,26,38), and mitigation of negative effects from staff turnover, particularly among physicians (10). Key SS components such as training, mentorship, on-the-job training and staff recognition were instrumental in fostering these improvements (10, 24, 34). Interestingly, even when Component 4 of the WHO SS guidelines—focused on mentoring and coaching—was excluded, 8 out of 10 studies still reported positive outcomes. Of these, eight measured effectiveness through improved knowledge and skills, while two assessed changes in attitude (14, 20, 26). Notably, Study (14) reported ineffective results, suggesting that Component 4 may play a critical role in bridging gaps in attitude-related outcomes. Beyond SS processes, external factors such as logistics, incentives, and salary also significantly influenced attitudinal behaviour (34). Moreover, improved attitudes were observed not only among supervisees but also among supervisors (8). However, challenges such as lack of performance-based incentives and logistical constraints negatively impacted supervisors’ motivation and ability to conduct SS, leading to reduced supervision frequency.

The second role of SS in primary care performance is its ability to **enhance health workers’ knowledge and skills** (11, 17, 22, 36). SS contributed to improved knowledge and skills among health workers, demonstrated by improved knowledge manifested in health workers’ ability to transfer knowledge to clients and/or fellow health workers (11), improved disease case definition and case management (17), and improved community health workers’ ability to serve their catchment population (22). While improved skills in primary care performance were manifested by supervisees’ improved ability to appropriately assess and diagnose conditions like pneumonia and dehydration (12, 13, 16, 20, 22). This is mainly linked to the third positive role of supportive supervision on primary health care performance.

The third role of supportive supervision on PHC performance is its ability to improve patient outcomes (27, 29, 30, 37). Improved patient outcome was seen as a result of improved clinical practice and improved service provision (12, 17, 26, 27, 30, 33, 36, 37). Improved service provision resulted from support and training for CHWs and supervisors’ new skills, addressing fears of failing & establishing operational systems to address inefficiencies. Direct employment of CHWs and increased stipends further motivated and integrated them into local PHC teams. However, the nurse mentor could not establish a collaboration with local structures, while excessive evaluation anxiety among Rwandan PHC providers posed challenges in establishing a supervisory relationship appropriate for generating professional development among providers. Madede et al. 2017

The third role of supportive supervision on PHC performance is its ability to improve patient outcomes (25, 27, 29, 30, 37). Improved patient outcome was seen because of improved clinical practice and improved service provision (12, 17, 26, 27, 30, 33, 36, 37). Improved service provision resulted from support and training for CHWs and supervisors’ new skills, addressing fears of failing & establishing operational systems to address inefficiencies. Direct employment of CHWs and an increase in their stipend added to their motivation and integration into the local PHC team. However, the nurse mentor could not establish a collaboration with local structures, while excessive evaluation anxiety among Rwandan PHC providers posed challenges in establishing a supervisory relationship appropriate for generating professional development among providers.

### Limitation of the review

Limitations to the study included the restriction to only include English articles, which may have potentially excluded some relevant studies that were written in other languages. Additionally, despite a thorough search, some relevant studies may have been missing in the process.

## CONCLUSION

The systematic review identified multiple approaches to conducting SS and that these mechanisms yielded positive, negative and inconclusive results, thus supporting evidence on inconsistencies in what constitutes an effective mechanism of supportive supervision in primary health care in the LMIC. This review looks at the following key information: A mechanism has the possibility to yield effective, ineffective or inconclusive results, as there are multiple factors that affect outcomes. Multiple external factors such as time, consistency, incentives, material and human resources influence SS outcomes both negatively and positively. The six parameters discussed as a WHO mechanism of SS can be easily used. There is a need to understand if on-the-job training and mentoring may not be key in improving health outcomes and improved service provision. Feedback, on-the-job training and development of action items were steps that were missed by the supervisors.

## Contributors

ANN conceptualized the study, designing the methods and had access to all data. FC compiled and executed the search. ANN and FC screened the articles and curated data under the supervision of LWD and EU. ANN and FC performed the data collection and critical appraisal under the supervision of LWD and EU. ANN drafted the paper with reviews and edits from all authors for the final draft. All authors take responsibility for the submission and content of the paper.

## Patient and public involvement

Patients and/or the public were not involved in the design, conduct, reporting, dissemination plans of this research.

## Ethics approval

Ethical review and approval was done by COMREC reference number **P.07/23-0141**.

## Provenance and peer review

Not commissioned; externally peer reviewed.

## Data availability statement

Data are available on reasonable request. The data collected for this review is available from the corresponding author on request.

Joanna Briggs Institute (JBI). (2024). *Critical appraisal tools*. JBI. https://jbi.global/critical-appraisal-tools

